# Ideology, policy decision-making and environmental impact in the face of the Coronavirus pandemic in the US

**DOI:** 10.1101/2021.09.01.21262952

**Authors:** Juan Prieto-Rodríguez, Rafael Salas, Douglas Noonan, Francisco Cabeza-Martinez, Javier Ramos-Gutiérrez

## Abstract

Covid-19 pandemic was a challenge for the health systems of many countries. It altered people’s way of life and shocked the world economy. In the United States, political ideology has clashed with the fight against the pandemic. President Trump’s denial prevailed despite the warnings from the WHO and scientists who alerted of the seriousness of the situation. Despite this, some state governments did not remain passive in the absence of federal government measures, and passed laws restricting mobility (lockdowns). Consequently, the political polarity was accentuated. On the one hand, the defenders of more severe public health measures and, on the other, the advocates of individual rights and freedom above any other consideration. In this study, we analyze whether political partisanship and the political ideology has influenced the way Covid-19 was handled at the outbreak. Specifically, we analyze by using a Diff-in-Diff model, whether the ideology of each state, measure at three levels, affected the decrease in the NO2 levels observed after the pandemic outbreak in the US. We distinguish three alternative post-Covid periods and results show that the State ideology has a robust negative impact on the NO2 levels. There is an important difference between Democratic and Republican states, not just in the scope and following-up of the mobility and activity restrictions, but also in the speed they implemented them.

## Introduction

The Covid-19 pandemic has been a challenge for many countries’ health systems, causing the death of thousands of people around the world. It has altered people’s way of life and shocked the world economy. The impact of Covid-19 has been anything but random, however, affecting different social groups differently (*1*). In the United States, political ideology has clashed with the fight against the pandemic (*2*). During the outbreak of the pandemic, President Donald Trump’s denial prevailed over the WHO warnings and scientists who warned of the seriousness of the situation (*3*). Despite this, state governments did not remain passive, and they issued executive orders restricting activities (lockdowns). Consequently, political polarization was accentuated, pitting defenders of more severe public health measures against advocates of individual rights and freedom. The states’ heterogeneous responses resulted in different behavioral responses in terms of mobility and economic activity that, in turn, led to significantly different trends in pollution. In this study, we analyze whether the political ideology of each state governor and, therefore, their voters influenced the decreases in NO_2_ levels or, on the contrary, did not undergo any change.

These drops are likely caused by restrictions on individual (recreational and retail) mobility and economic activity as happened in other countries (*4-5*) which, in turn, are linked to legal requirements due to lockdown policies but, also, to precautionary measures taken voluntarily by the public before or even after lockdowns enforcement (*6*-*7*). Both causes, however, may be related to ideology and, ultimately, to the electorate ideology. However, ideology has a dual channel to influence mobility, activity and, ultimately, pollution. First, ideology has a direct pathway to mobility/activity and therefore pollution, through people’s behavior, that may depend on the confidence in political leaders (*8*). But, second, it also has an indirect pathway by causing policy, which in turn affects behavior (*6*) and, hence, mobility/activity and finally pollution, through executive orders. The two pathways for ideology to affect mobility/activity and pollution allows us to identify whether policy is driving the results, or it is just the ideology that directly affects activity and pollution, or a combination of the two. In order to assess this double path, ideology has been measured by using three alternative variables: governor’s party affiliation, Trump’s share of the votes in 2016 presidential election at county and state level.

In this sense this research constitutes additional evidence, complementary to Neelon et al. (*2*), about the importance of ideology and partisanship in the way the US faced and managed the Covid-19 outbreak. Declining pollutant levels also have other major effects on people’s health. In a study by Caiazzo et al. (*9*), they concluded that 200,000 people die prematurely in the US due to pollution, reducing the life expectancy of the affected people by a decade. Furthermore, Chossière et al. (*10*) find that cuts in NO_2_ concentrations associated to the Covid-19 resulted in about 32,000 prevented premature deaths, including about 21,000 in China. But NO_2_ cuts affect not only health but also the environment, as NO_2_ generates acid rain (*11*).

In recent years, the two main political parties in the United States have taken divergent policy positions. For example, the Democratic party has emphasized stronger protections for the environment while the Republican party has prioritized individual freedoms and reductions in environmental regulation. Different opinion polls, such as the one carried out by the Pew Research Center (*12*), show a large gap in the environmental concerns of the members and voters of the two main parties. With the arrival of Donald Trump to the White House, he has favored energy deregulation and the use of fossil fuels. The new regulation replaced the Clean Power Plan with the Affordable Clean Energy (ACE) rule, giving states more power to decide on emission limits for their plants and eliminating many of the US Environmental Protection Agency’s (EPA) oversight powers. Similarly, at the state level, these divergent environmental positions are increasingly clear, with several Democratic states pushing very ambitious laws, such as the 2019 New York bill that aims to reduce net greenhouse gas emissions to zero by 2050 (*13, 14*).

The high contagion rate of Covid-19 has led policymakers to take measures to restrict individual mobility to prevent its spread, which has caused sharp drops in economic activity and the pollution it creates. In response to the crisis brought on by the Covid-19 crisis, politicians and their appointed officials weigh the health of citizens, capacity of healthcare systems, and the strength of their economies. The pandemic offers an opportunity to examine how different priorities associated with political partisanship and ideology manifest in different environmental outcomes.

In the US, the first cases of Covid-19 were registered in late February 2020 and, since then, the public health crisis expanded rapidly, reaching more than 33.5 million infected and almost 600 thousand deaths on May 12, 2021 (Centers for Disease Control and Prevention). This growth may be related to the position of Donald Trump, who was very skeptical of the scientific evidence on this disease, even recommending the intake of bleach to eliminate the virus (*15*) and who, consequently, refuses to take severe measures that may affect the economy and businesses. Management of the pandemic by Trump’s administration prompted the New England Journal of Medicine to publish an editorial criticizing it, stating that “they have taken a crisis and turned it into a tragedy” (*3*).

Not all policymaking authority, however, rests with the nation’s president. In the United States, governors, as representatives of the executive branch of each state, maintain high levels of autonomy, while the legislative branch at the state level (usually with a bicameral structure) has significant legislative powers. However, very important differences were observed between Democratic and GOP states. For instance, the first orders requiring the use of face masks were issued in April 2020 by seven different states: Connecticut, Delaware, Hawaii, Illinois, Maryland, New Jersey and New York. Only one (Maryland) has a Republican governor. Five states passed similar orders in June 2020, all of their governors were Democrats. On the other hand, of 14 states that did not approve orders for the imposition of face masks, twelve had Republican governors. Moreover, Arkansas, Iowa, Nebraska, North Dakota, Oklahoma, South Dakota, Utah and Wyoming did not pass a general state “stay-at-home order;” they all had Republican governors.

Since the appearance of Covid-19 can be considered an unexpected event, it is assumed that the distribution by political parties of the different governors and the Trump’s share of the votes in 2016 presidential election are orthogonal to the emergence of the disease (conditional on public health drivers). Consequently, it is assumed that the emergence of Covid-19 is a quasi-natural experiment where the air quality stations included in the sample are considered treated or a control depending on the party of the governor of the state in which they are located. Based on this assumption, we use data from the EPA to check whether the observed drops in the levels of NO_2_ in different cities in the US since the outbreak of the Covid pandemic have had any relationship with the political sign of the state governor.

This research uses difference-in-difference models (Diff-in-Diff) comparing the evolution of the levels of NO_2_ from the beginning of 2018 to July 2020. We expect that the political affiliation of each state and the political party of their governors have affected the evolution of pollution levels differently. In particular, if Republican voters and Republican governors have advocated for the defense of individual liberty, we expect the falls in the levels of the polluting gases to be lower than those observed in the Democratic states.

## Results

We observed a significant reduction of NO_2_ levels after the Covid-19 outbreak in the United States between February and March 2020. Figure 1 shows 1-week moving averages of NO_2_ levels by governor’s political party, using the date of the first case of community spread on February 26, 2020 to differentiate the period previous to the outbreak of the pandemic in US to the post-pandemic period (hereafter referred to as the “pre” and “post” periods). Figure 1 is only a first approximation to understand what effect the political parties in the US have had on the levels of pollution and contamination after the outbreak of the Covid-19 pandemic, mainly related with the toughness and speed of their confinement measures. During the first weeks of the Covid-19 pandemic in US, important decreases in NO_2_ took place independently of the state governor’s party. Moreover, Democratic states had higher levels of both pollutants before the lockdown that almost converged to Republican states’ levels after the first wave, reflecting, thus, larger cuts in Democratic states. A common characteristic of many polluting gases, including NO_2_, is their origin. They are present in cities and industrial areas due mainly to combustion processes (power plants, motor vehicles, heating, etc.) (*16*). This explains the differences in average levels of pollution of some states in relation to others, as their percentages of urban population, industry and chemical sectors differ. In general, the more urban and industrialized states lead to higher concentrations of NO_2_. This implies that any statistical model to explain NO_2_ levels must include air stations fixed effects to control for these differences.

**Figure 1:**
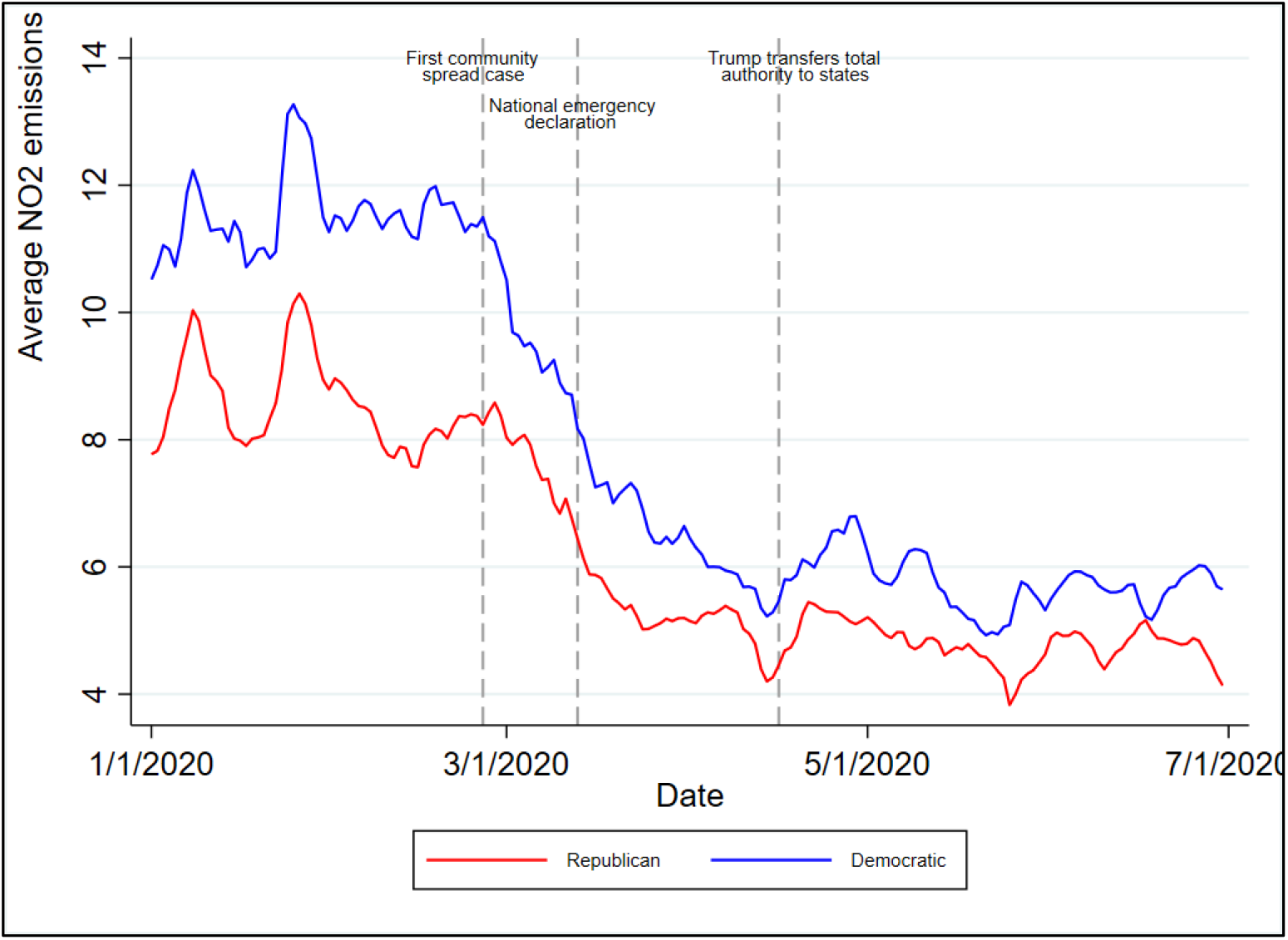
Evolution of NO_2_ levels in Republican and Democratic states.

Regardless of the intense drop in NO_2_ at the end of the first quarter of 2020, Figure 2 shows that similar drops are also observed in previous years around the same weeks of the year. Using weekly moving averages, between February 26 (first case of community spread) and April 16, 2020 (when President Trump proclaimed a transfer of responsibility on how to restart shuttered activity to state governors), there was a 53 percent reduction in the cross-station average NO_2_ level. By contrast, the cuts corresponding to the same dates in 2019 and 2018 were just 27 and 10 percent for NO_2_ in these two years. This suggests that the observed reduction in 2020, compared with the same days of the two previous years, was much larger for this pollutant. But it is still important to identify whether the 2020 pollutant reductions were larger than usual due to the economic activity decline associated to the Covid-19 lockdown and, therefore, statistically different to the falls observed previously. To do this, we have included year-month fixed effects to capture both longer-run trends in and seasonal effects of air quality.

**Figure 2:**
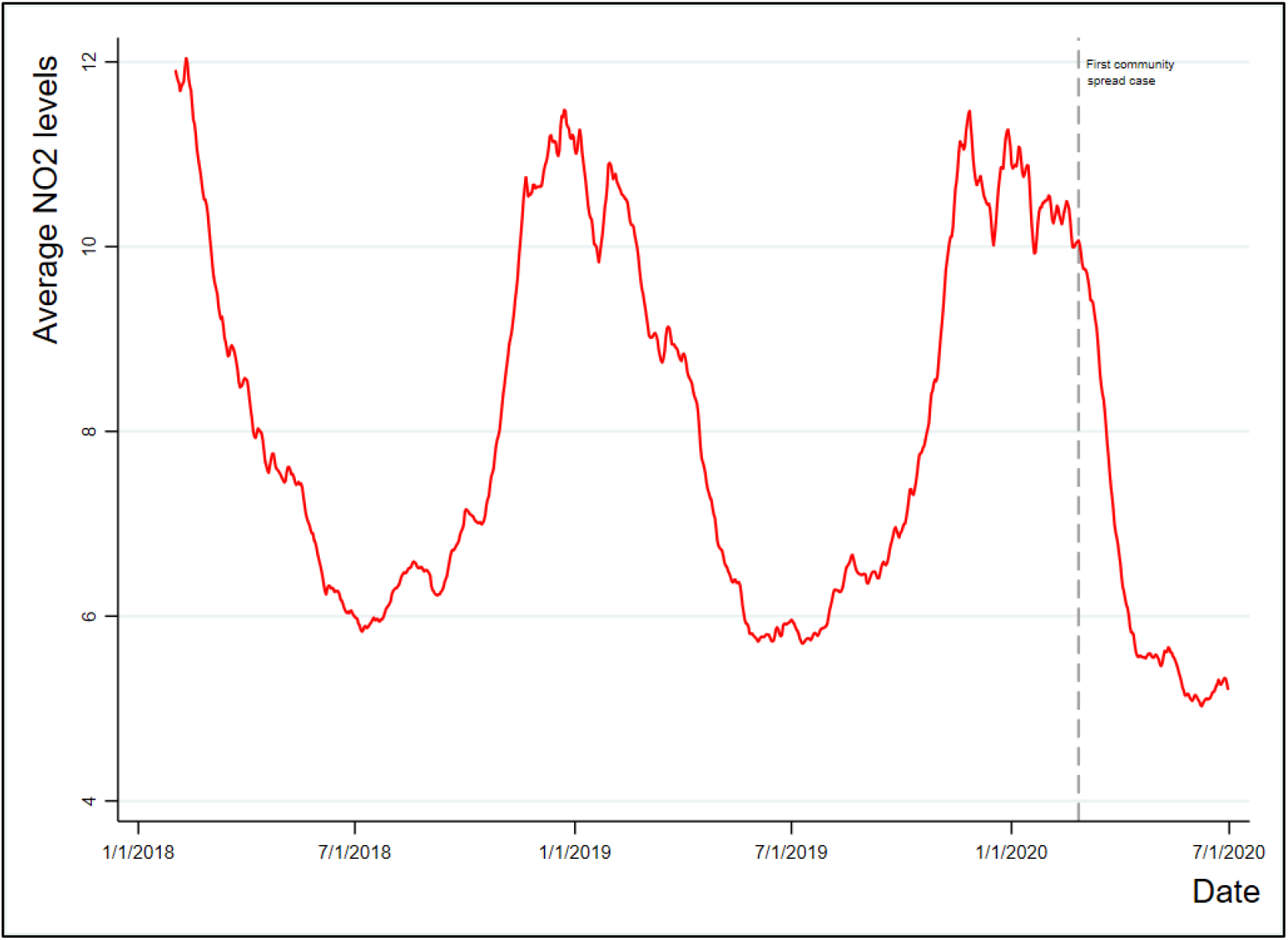
Evolution of average NO_2_ levels.

Additionally, these ups and downs are related to the divergent evolution of NO_2_ levels in Democratic and Republican states through the year, which, according to Figure 3, may present a stable pattern in recent years. As the difference is usually negative, the average levels of NO_2_ in Democratic states are higher than in the Republicans. Furthermore, at the end of winter and early spring, a huge drop in these levels is observed, which is associated with a significant increase in the difference between the two types of states. Moreover, if the reduction in the levels of these pollutants occurs simultaneously with the increase in the difference between the states of both political affiliations, this should be due to a more rapid and pronounced drop in the levels of NO_2_ in the Democratic states compared to Republicans during these months. These differences may be related with the heterogeneous geographical distribution of Republican and Democratic states with different climate regimes and seasonal climate fluctuations. Therefore, it is crucial to identify whether the increase in the difference between states during the first half of 2020 is comparable to the observed increase in 2018 and 2019 or whether part of it can be associated with the drop in activity due to the lockdown. Hence, it is necessary to control for regional seasonal differences to verify if there are statistically significant differences when comparing states of different political affiliations in 2020 compared to previous years.

**Figure 3.**
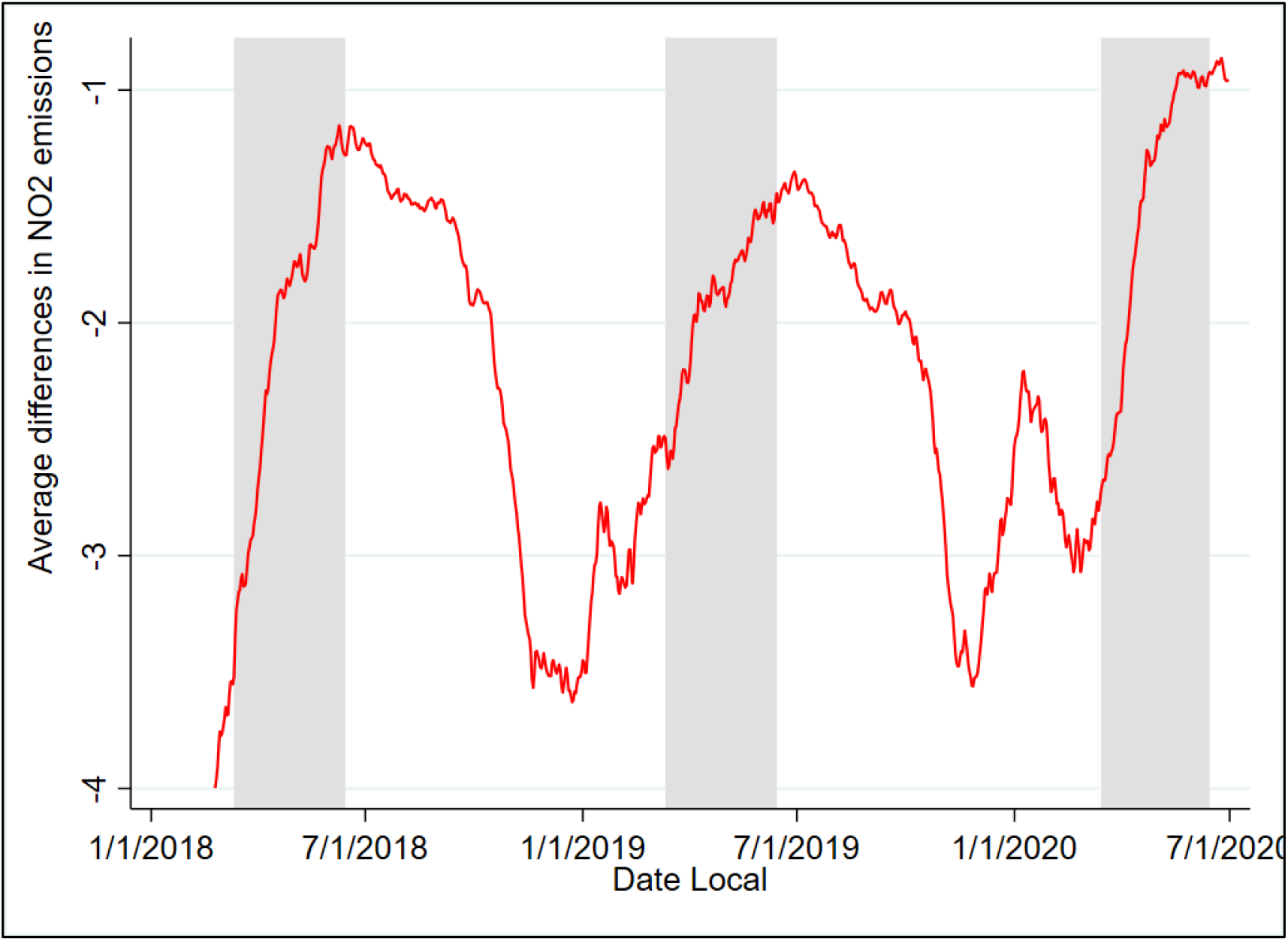
Differences in Republican-Democratic NO_2_ average levels.

Finally, since in the case of Covid-19, the type and timing of interventions affected the course of the pandemic (*8*), we used Diff-in-Diff models controlling for three different periods: *P1*, observations from February 26 to March 12 (day before President Trump declared a nationwide emergency); *P2*, from March 13 to April 15, 2020 (day before President Trump transferred responsibility on how to restart shuttered activity to state governors) and *P3* since April 16, 2020 to June 30, the last day with consolidated data on NO_2_ reported by the EPA.

Finally, ideology has been measured by using three alternative variables: governor’s party affiliation (*RepG*), Trump’s share of the votes in 2016 presidential election at, first, county level (*TrumpC*) and, second, state level (*TrumpS*). These three variables emphasis different aspects of the potential link between ideology and the impact on how Covid-19 pandemic was handled. The first variable could be a better proxy for the indirect effect through the policies enacted by elected officials. Since NO_2_ concentrations are measured locally, the degree of county’s Republican-ness could be a good signal of the direct impact of ideology on pollution through the individual behavior of voters living in the neighborhood of the air quality stations. Finally, the state Republican-ness could be a mix of both, the direct and the indirect impacts. It captures the degree to which governor’s policy is modulated by social pressure but also the percentage of voters of each party might have an impact. However, the ideological variables are invariant in time by air quality measurement station and, therefore, their effect will be captured by the estimated fixed effects together with the effects of the rest of the fixed factors. What is relevant is the differential effect of ideology during the post Covid-19 outbreak periods. Hence, the interaction terms between these three ideological variables and the three periods after the Covid outbreak were defined. The coefficients associated to these variables will be the Diff-in-Diff estimators of the NO_2_ drops due to the ideology effect.

The correlations between these interaction terms are significant and very high (Table 1). Therefore, it can be assumed that they similarly capture the effect of ideology, but not in exactly the same way. The interaction terms between the percentage of Trump voters by county (*P*_*i*_*\*TrumpC*) and state (*P*_*i*_*\*TrumpS*), for *i=1,2,3*; have correlation coefficients of around 94 percent, i.e. differences between these two variables are very small and probably linked to the larger degree of the direct effect capture by the percentage of Trump voters by county.

**Table 1.** Within periods correlation between the ideological variables.

|  | *P1\*RepG* | *P1\*TrumpC* | *P2\*RepG* | *P2\*TrumpC* | *P3\*RepG* | *P3\*TrumpC* |
| --- | --- | --- | --- | --- | --- | --- |
| *P1\*TrumpC* | 0.78 | 1 |  |  |  |  |
| *P1\*TrumpS* | 0.75 | 0.94 |  |  |  |  |
| *P2\*TrumpC* |  |  | 0.78 | 1 |  |  |
| *P2\*TrumpS* |  |  | 0.75 | 0.94 |  |  |
| *P3\*TrumpC* |  |  |  |  | 0.77 | 1 |
| *P3\*TrumpS* |  |  |  |  | 0.74 | 0.94 |

Table 2 reports the results for different combinations of the Diff-in-Diff estimators for NO_2_ levels, measured in parts per billion. All estimations include station fixed effects, which control for all time-invariant characteristics of the monitoring sites, and help to achieve a fairly high explanatory power of the models, measured by the R^2^. Other time and regional fixed effects already mentioned were also considered in addition to a set of weather and bank holidays variables.

**Table 2.**
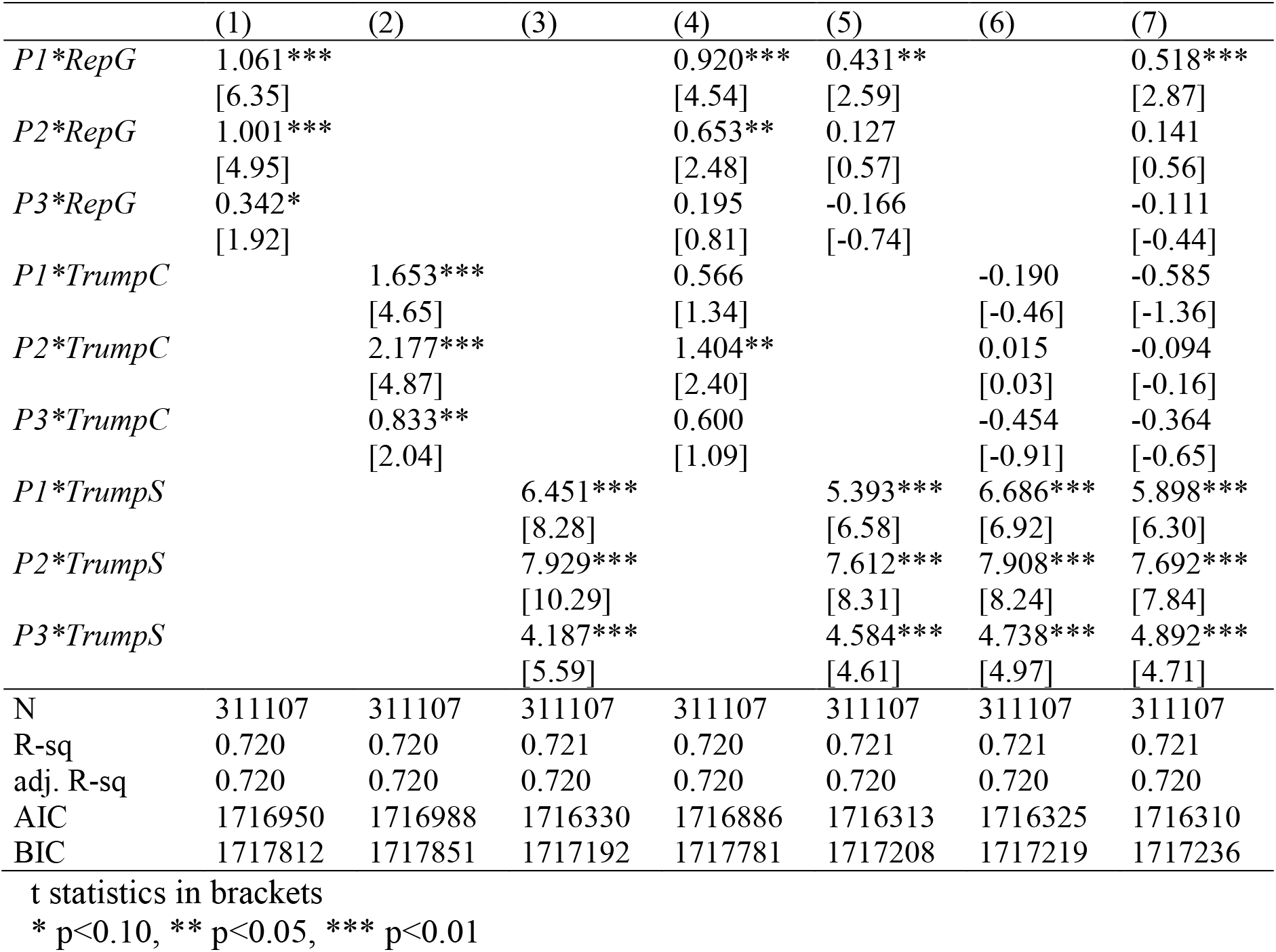
Diff-in-Diff estimators of the additional drops in NO_2_ in Democratic states.

|  | (1) | (2) | (3) | (4) | (5) | (6) | (7) |
| --- | --- | --- | --- | --- | --- | --- | --- |
| *P1\*RepG* | 1.061***<br>[6.35] |  |  | 0.920***<br>[4.54] | 0.431**<br>[2.59] |  | 0.518***<br>[2.87] |
| *P2\*RepG* | 1.001***<br>[4.95] |  |  | 0.653**<br>[2.48] | 0.127<br>[0.57] |  | 0.141<br>[0.56] |
| *P3\*RepG* | 0.342*<br>[1.92] |  |  | 0.195<br>[0.81] | -0.166<br>[-0.74] |  | -0.111<br>[-0.44] |
| *P1\*TrumpC* |  | 1.653***<br>[4.65] |  | 0.566<br>[1.34] |  | -0.190<br>[-0.46] | -0.585<br>[-1.36] |
| *P2\*TrumpC* |  | 2.177***<br>[4.87] |  | 1.404**<br>[2.40] |  | 0.015<br>[0.03] | -0.094<br>[-0.16] |
| *P3\*TrumpC* |  | 0.833**<br>[2.04] |  | 0.600<br>[1.09] |  | -0.454<br>[-0.91] | -0.364<br>[-0.65] |
| *P1\*TrumpS* |  |  | 6.451***<br>[8.28] |  | 5.393***<br>[6.58] | 6.686***<br>[6.92] | 5.898***<br>[6.30] |
| *P2\*TrumpS* |  |  | 7.929***<br>[10.29] |  | 7.612***<br>[8.31] | 7.908***<br>[8.24] | 7.692***<br>[7.84] |
| *P3\*TrumpS* |  |  | 4.187***<br>[5.59] |  | 4.584***<br>[4.61] | 4.738***<br>[4.97] | 4.892***<br>[4.71] |
| N | 311107 | 311107 | 311107 | 311107 | 311107 | 311107 | 311107 |
| R-sq | 0.720 | 0.720 | 0.721 | 0.720 | 0.721 | 0.721 | 0.721 |
| adj. R-sq | 0.720 | 0.720 | 0.720 | 0.720 | 0.720 | 0.720 | 0.720 |
| AIC | 1716950 | 1716988 | 1716330 | 1716886 | 1716313 | 1716325 | 1716310 |
| BIC | 1717812 | 1717851 | 1717192 | 1717781 | 1717208 | 1717219 | 1717236 |

t statistics in brackets
\* p<0.10, \*\* p<0.05, \*\*\* p<0.01

Defined as the interaction terms between the three ideological variables and the three sub-periods after the Covid-19 outbreak, all the estimated coefficients for the Diff-in-Diff effects are positive when using just one set of variables (columns 1 to 3), and all significantly different from zero. That means the Republican states had significantly smaller cuts in the NO_2_ levels, regardless of how the COVID-19 starting point is defined or how the ideology effect is measured. Moreover, according to the Akaike Information Criterion, the interaction terms defined from Trump’s share of votes by state (*TrumpS*) is the best way of introducing ideology in the empirical model, being the governors’ party the second-best alternative and Trumps’ votes by county the third. In columns 4 to 6, the interactions terms are introduced by pairs, obtaining the best results, again, when ideology is proxied by *TrumpS* combined with *RepG*. When the two voting variables are jointly considered, no interaction term defined using *TrumpC* is significant while the three with *TrumpS* are. This is an insight that voters’ ideology may have an impact on NO_2_ but the indirect effect is more relevant than the direct one, more closely linked to *TrumpC*. Finally, the best alternative according to the AIC is estimation in column 7, including the three set of interaction terms simultaneously. Results are consistent with the previous models and again *P*_*i*_*\*TrumpS* variables are the most significant. Still, *P*_*i*_*\*TrumpC* variables are not significant. Having a Republican governor only had a significant extra effect on top of the State share of Trump’s voter during P1, reinforcing the ideology indirect effect, especially at the very beginning of the pandemic.

As the three post-outbreak period variables do not overlap, estimated effects refer to the Republican lower reduction in that particular period, compared to the pre-Covid situation. Also, the difference between the Democratic and Republican states in the NO_2_ drops narrowed after April 16, since the smaller cut in Republican states in the NO_2_ levels was lower than in previous weeks but, still, significant. Therefore, there was an important difference between Democratic and Republican states not just in the scope and following-up of their mobility and activity restrictions but also in the speed at which they took them. This result seems to be very robust, since the estimated difference is robust to the different specifications of the ideology variable. Also, these results are robust to different specifications of the post-outbreak periods and to different definitions of the sample period.

Regarding the impact of the rest of independent variables, Table 3 reports the whole set of estimated coefficients for the specifications in columns 3, 5 and 7 of Table 2. As expected, during rainy days we tend to observe lower levels of pollutants, but there is a certain degree of saturation of this effect on atmospheric pollutants (*17*). Dew point temperature is negatively linked to NO_2_ levels (*6*). Additionally, we estimate a non-linear effect for the atmospheric pressure, as in Borge et al. (*18*) or Roberts–Semple *et al*. (*19*). Given the estimated coefficients and the shape of the curvature, the positive relationship between pressure and NO_2_ predominates. Therefore, the higher the sea-level pressure values, the higher the expected levels of NO_2_, though this effect moderates when pressure is above 1025 hPa. Wind speed presents a negative relationship with NO_2_ levels, but with a decreasing rate; as wind speed reduces pollution levels, but the higher the speed, the lower its marginal effect.

**Table 3.** NO_2_ regression models.

|  | Model 1 | Model 2 | Model 3 |
| --- | --- | --- | --- |
| P1*RepG | -- | 0.431**<br>[2.59] | 0.518***<br>[2.87] |
| P2*RepG | -- | 0.127<br>[0.57] | 0.141<br>[0.56] |
| P3*RepG | -- | -0.166<br>[-0.74] | -0.111<br>[-0.44] |
| P1*TrumpC | -- | -- | -0.585<br>[-1.36] |
| P2*TrumpC | -- | -- | -0.094<br>[-0.16] |
| P3*TrumpC | -- | -- | -0.364<br>[-0.65] |
| P1*TrumpS | 6.451***<br>[8.28] | 5.393***<br>[6.58] | 5.898***<br>[6.30] |
| P2*TrumpS | 7.929***<br>[10.29] | 7.612***<br>[8.31] | 7.692***<br>[7.84] |
| P3*TrumpS | 4.187***<br>[5.59] | 4.584***<br>[4.61] | 4.892***<br>[4.71] |
| Precipitation in mm | 2.7e-04<br>[0.04] | 2.7e-04<br>[0.04] | 2.8e-04<br>[0.04] |
| Precipitation in mm squared | 2.8e-04***<br>[2.77] | 2.8e-04***<br>[2.77] | 2.8e-04***<br>[2.77] |
| Dew point in Celsius | -0.219***<br>[-15.10] | -0.219***<br>[-15.13] | -0.219***<br>[-15.12] |
| Dew point in Celsius squared | -0.003***<br>[-6.38] | -0.003***<br>[-6.39] | -0.003***<br>[-6.38] |
| Pressure in Pa | 0.022***<br>[3.29] | 0.022***<br>[3.30] | 0.022***<br>[3.29] |
| Pressure in Pa squared | -1.1e-07***<br>[-3.25] | -1.1e-07***<br>[-3.25] | -1.1e-07***<br>[-3.24] |
| Wind intensity in m/s | -3.051***<br>[-23.47] | -3.050***<br>[-23.47] | -3.050***<br>[-23.48] |
| Wind intensity in m/s squared | 0.195***<br>[14.67] | 0.195***<br>[14.67] | 0.195***<br>[14.67] |
| Temperature in Celsius | 0.208***<br>[11.85] | 0.208***<br>[11.88] | 0.208***<br>[11.87] |
| Saturday | -1.491***<br>[-19.84] | -1.491***<br>[-19.84] | -1.491***<br>[-19.84] |
| Sunday | -2.332***<br>[-22.82] | -2.332***<br>[-22.82] | -2.332***<br>[-22.82] |
| New Year's Day | -3.413***<br>[-19.42] | -3.413***<br>[-19.42] | -3.413***<br>[-19.42] |
| Martin Luther King Jr. Day | -1.149***<br>[-8.39] | -1.148***<br>[-8.39] | -1.148***<br>[-8.39] |
| Presidents day | -1.373***<br>[-11.11] | -1.372***<br>[-11.11] | -1.372***<br>[-11.11] |
| Memorial day | -2.463***<br>[-17.45] | -2.463***<br>[-17.45] | -2.464***<br>[-17.45] |
| Fourth of July | -1.730***<br>[-13.24] | -1.730***<br>[-13.24] | -1.730***<br>[-13.24] |
| Labor Day | -2.881***<br>[-17.41] | -2.881***<br>[-17.41] | -2.882***<br>[-17.41] |
| Columbus Day | -1.044***<br>[-8.38] | -1.044***<br>[-8.38] | -1.044***<br>[-8.38] |
| Veterans Day | -0.478***<br>[-3.01] | -0.477***<br>[-3.01] | -0.477***<br>[-3.01] |
| Thanksgiving | -4.267***<br>[-20.00] | -4.267***<br>[-20.00] | -4.266***<br>[-20.00] |
| day after Thanksgiving | -2.743***<br>[-17.16] | -2.743***<br>[-17.16] | -2.742***<br>[-17.17] |
| Christmas | -4.128***<br>[-19.24] | -4.129***<br>[-19.24] | -4.128***<br>[-19.24] |
| Station fixed effects | YES | YES | YES |
| Region-quarter fixed effects | YES | YES | YES |
| Year-month fixed effects | YES | YES | YES |
| N | 311107 | 311107 | 311107 |
| R-sq | 0.721 | 0.721 | 0.721 |
| AIC | 1716329.9 | 1716313.4 | 1716309.8 |
| BIC | 1717192.4 | 1717207.8 | 1717236.1 |

t statistics in brackets. \* p<0.10, \*\* p<0.05, \*\*\* p<0.01

Daily average temperatures show a significant positive sign. In sum, daily temperature, high pressure, low precipitation and weak winds are associated with higher levels of NO_2_, since this combination favors stable lower atmosphere that effectively prevents vertical dispersion, and the absence of precipitation or winds to wipe pollution away. Finally, not surprisingly, NO_2_ levels are generally lower during the weekends, especially Sundays, and the bank holidays of Thanksgiving and Christmas had the largest impact on NO_2_ reduction.

In order to study how pollution reductions in Democratic states relative to Republican-ruled ones are geographically distributed, a model including station fixed effects for each subperiod was also estimated. Then, the average of the station fixed effects by subperiod and state were computed. Results, in Figure 4, show the average NO_2_ reductions and colors are differentiated according to values higher or lower than the median. Yellow represents lower reductions and green denotes larger ones. Yellow is more common in southern and central regions, usually with larger Republican voter shares and Republican governors; while green represents higher reductions associated more frequently with states ruled by Democratic governors.

**Figure 4.**
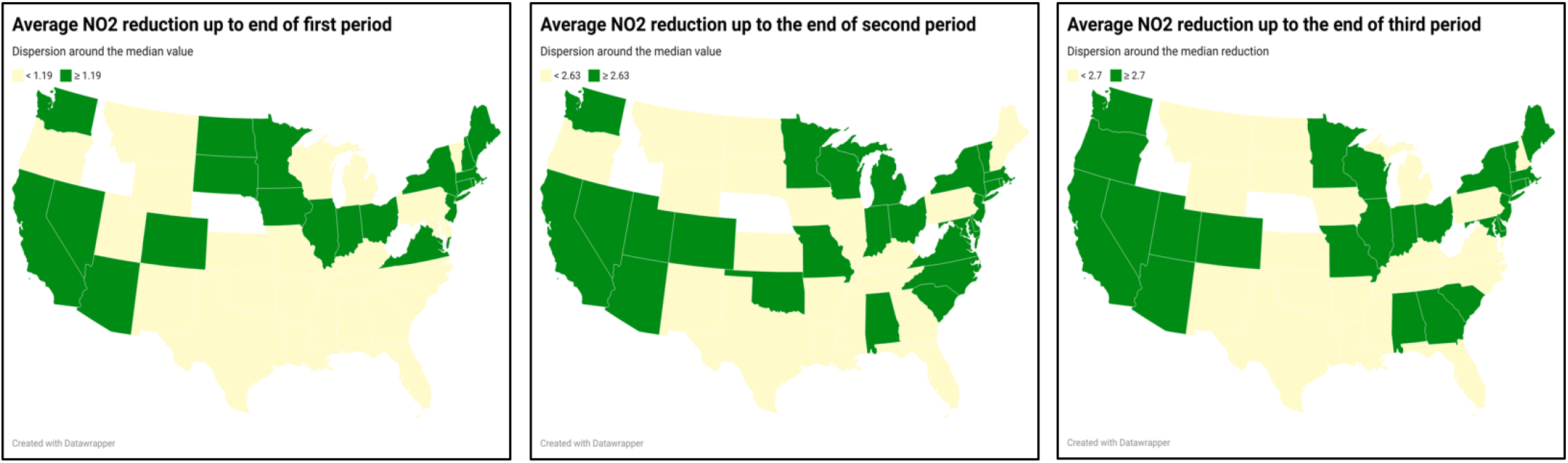
Average pollutants reduction in the three periods.

Table 4 shows the t-tests on the mean differences of the fixed effects of the air quality stations, separately for the three subperiods, comparing the states governed by Democratic governors with those under Republican governors, assuming an unequal variance of both samples. A negative value of the difference implies a larger reduction in the Democratic states, which occurs in all three periods. Very similar mean differences of -1.02 and -1.03 were observed at the first and second periods, respectively, both significant at 0.1%. NO_2_ difference during the third subperiod was -0.38, significant at 10%.

**Table 4.**
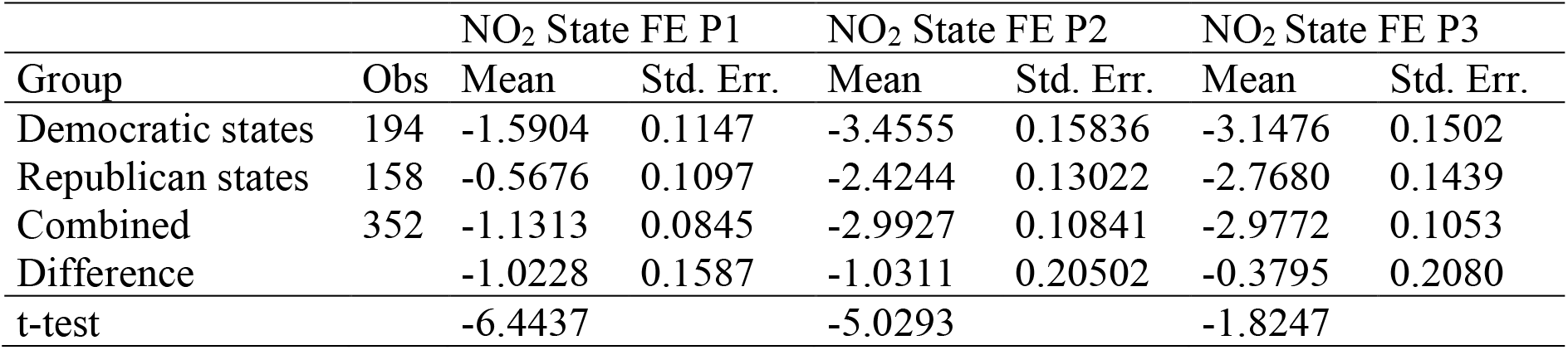
T-test in mean differences between Democratic and Republican states.

| Group | Obs | NO<sub>2</sub> State FE P1 |  | NO<sub>2</sub> State FE P2 |  | NO<sub>2</sub> State FE P3 |  |
| --- | --- | --- | --- | --- | --- | --- | --- |
|  |  | Mean | Std. Err. | Mean | Std. Err. | Mean | Std. Err. |
| Democratic states | 194 | -1.5904 | 0.1147 | -3.4555 | 0.15836 | -3.1476 | 0.1502 |
| Republican states | 158 | -0.5676 | 0.1097 | -2.4244 | 0.13022 | -2.7680 | 0.1439 |
| Combined | 352 | -1.1313 | 0.0845 | -2.9927 | 0.10841 | -2.9772 | 0.1053 |
| Difference |  | -1.0228 | 0.1587 | -1.0311 | 0.20502 | -0.3795 | 0.2080 |
| t-test |  | -6.4437 |  | -5.0293 |  | -1.8247 |  |

The fall in NO_2_ levels between February 26 and March 12 in the Democratic states is slightly lower than the NO_2_ reduction associated with the celebration of the Fourth of July, that is, as if any working day of the first week of March 2020 would have become the Fourth of July in terms of NO_2_ pollution levels. However, in the Republican states, this cut was only marginally larger than that of Veterans Day. Moreover, the declines, from March 13 to the end of June 2020, were similar to NO_2_ drops on New Year’s Day in Democratic states but similar to those on Memorial Day or Labor Day in the Republican states. Likewise, the differences in the average drops for this pollutant between these two types of states are smaller after April 16, 2020 (*P3*), that is, the downward adjustments seem to converge at the end of the period analyzed, but are still statistically greater in the Democratic states.

## Discussion

Donald Trump, a vocal Republican defender of individual liberties and the country’s most populist wing, has become one of the pandemic’s biggest deniers. His inactivity may be one of the causes of the rapid spread of the disease throughout the US. On March 13, given the worsening of the situation, he declared a national emergency (Trump, 2020).

However, his limited efforts to contain the pandemic received considerable criticism and within days the country was already immersed in a high number of infections, reaching 100 infected per 1,000,000 inhabitants on March 22, 2020. Given the high numbers of infections and the first deaths, some state governments restricted mobility and issued state orders to “Stay Home, Stay Healthy”. For instance, on March 23, there were already eight states in lockdown: California (March 19), Illinois (March 21), New York (March 22), Connecticut, Louisiana, Ohio, Oregon and Washington. Out of these states, only Ohio’s governor was Republican. Therefore, the speed in taking anti-Covid-19 measures related to mobility or economic activity restrictions (the most effective measures before the vaccine was available) hewed closely to the governor’s political party.

Despite the unfavorable evolution of the pandemic, many citizens claimed their individual rights and freedoms by demonstrating throughout the country at the end of April. One of the largest demonstrations took place in Austin (Texas). Citizens claimed their right to work and freedom of movement throughout the national territory. Tensions rose over lost economic opportunities and freedoms due to lockdowns, risky behaviors and inadequate public health protections, and disputes over optimal policy responses (e.g., public health strategies, support for those suffering economic hardship). This context seems appropriate to analyze whether there have been systematic differences between the two dominant parties in the US when facing the Covid-19 pandemic.

This research has used difference-in-difference models comparing the evolution of NO_2_ levels from the beginning of 2018 to July 2020. We showed that the political affiliation of the voters of each state and the political party of their governors has affected the evolution of pollution levels differently. All the estimated coefficients for the DiD variables (interaction terms between the three alternative specifications of the ideology variable and three sub-periods after the Covid-19 outbreak considered) are positive and significantly different from zero when using just one set of variables. Given how these ideology variables were defined, this implies the Republican states had significantly smaller cuts in the NO_2_ levels, regardless of how the COVID-19 starting point is defined or how the ideology effect is measured. Moreover, Trump’s share of votes by state is the best way of introducing ideology in the empirical model. Therefore, voters’ ideology may have an impact on NO_2_ but the indirect effect is more relevant than the direct one. Finally, given our results, having a Democratic governor only had a significant extra effect on top of the State share of votes during the very beginning of the pandemic, reinforcing the ideology indirect effect at the earliest stage of Covid-19 outbreak. This is consistent with the prominent role played by some governors at the outbreak of the pandemic, *ceteris paribus* the distribution of voters. Finally, the difference between the Democratic and Republican states in the NO_2_ drops narrowed after April 16. Therefore, there was an important difference between Democratic and Republican states not just in the scope and following-up of their mobility and activity restrictions but also in the speed at which they took them. This result seems to be very robust, since the estimated difference is robust to the different specifications of the ideology variable.

These results are a clear example of how ideology is often not independent, as it should be, from scientific issues regarding public health matters. This should make us think about the best strategy to follow in the future. Clear protocols for leadership by specialized health personnel should be established in future pandemics. This should be a good course of action that isolates us from purely ideological issues that may not be efficient or equitable.

## Materials and Methods

### Weather data

In order to calibrate the model, we have included meteorological controls from the ERA5 gridded dataset. The ERA5 data are reanalysis data produced by the European Centre for Medium-Range Weather Forecasts (*20*) and provide hourly estimates of a large number of atmospheric, land and ocean climate variables, including the meteorological variables used in our analysis. In particular, we use data on temperature, pressure, wind speed and relative humidity. They were retrieved on an hourly basis for the 2018-2020 period, on a gridded 0.25° x 0.25° spatial resolution, for the selected area encompassing the 45 contiguous states. Data were converted into daily data for the analysis and imputed to the monitoring stations by interpolation consisting of the use of weights of the inverse square distance from each air monitoring station to the closest gridded nodes.

Originally, all weather data are hourly and converted to daily data. Temperature and dew point data are both converted to daily average from °K to °C. Wind intensity is expressed in average daily m/s. Sea level pressure is average daily in pascals and, finally, total precipitation is total daily mm.

### NO_2_ concentration data

The data comes from the EPA, a regulatory agency authorized by Congress to write regulations to implement laws that aim to protect health and the environment. The US has a network of more than 4,000 stations with air quality monitors spread throughout the country. These stations are owned and managed by state environmental agencies, which submit the collected observations of pollutant concentrations to the AQS (air quality system), the database of the US EPA. To carry out the empirical analysis of this work, data have been collected for NO_2_ levels, measured in parts per billion, for the years 2018, 2019 and 2020 until June 30, with the units of measurement being parts per billion. The EPA updates every six months (spring and autumn) the published files with the validated data on pollutants from each season, so they have not yet made them available to the public for the months corresponding to the second half of 2020.

### Ideology data

A practical manner to capture voter’s ideology is by three variables: governor’s party affiliation (*RepG*), Trump’s share of the votes in 2016 at the county (*TrumpC*) and the state (*TrumpS*) level presidential election. The idea is to control for the governor’s party affiliation, the county’s Republican-ness and the state’s Republican-ness and the influence voters may have had on the mobility and activity cuts or how these drops could be due to legal initiatives by the governors.

Trump’s share of the votes in 2016 at the state level is obtained from the U.S. Federal Election Commission website. Trump’s share of the votes in 2016 at the county level is obtained from the Election, COVID, and Demographic Data by County dataset from the Kaggle website (https://www.kaggle.com/etsc9287/2020-general-election-polls).

### Empirical model

To analyze the effects of the ideology of each state on the NO_2_ concentrations, we use the following difference in difference models that predict the concentrations of NO_2_. To account for area- and time-specific confounders and to identify the causal effects of governors’ political party, county- and state-level Republican-ness on pollutants levels, we use air station fixed-effects and year-by-month fixed-effects. Geographic fixed effects account for the possibility that different areas have varying baselines of NO_2_ levels. Temporal fixed effects account for national-level changes in these pollutant levels due to long-term changes or the general impact of the pandemic. Regression models use clustered standard errors on the air quality station to address potential serial correlation problems.

Formally, the models, estimated for NO_2_ levels, can be expressed as follows:

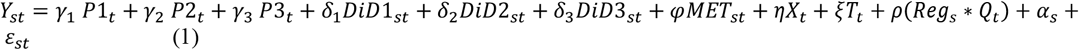

where *Y*_*st*_ is the pollutant variable for NO_2_, measured in parts by billion, and denotes daily average concentrations measured at the monitoring station *s* at time *t*; *P*1_*t*_ is a dummy variable taking value 1 between February 26 (first community spread case) and March 12, 2020 (day before President Trump declared a nationwide emergency) and zero otherwise; *P*2_*t*_ is a dummy taking value 1 between March 13 and April 15, 2020 (day before President Trump announced that state governors would be responsible for how to restart shuttered activity) and zero otherwise; and *P*3_*t*_, similarly, is a dummy variable that takes value 1 since April 16, 2020 to June 30, the last day with consolidated data on NO_2_ reported by the EPA. *DIDi*_*st*_ (*i*=1,2,3) is a vector defined by any of the *Pi*_*t*_*\*RepG*_*s*_, *Pi*_*t*_*\*TrumpC*_*s*_ and *Pi*_*t*_*\*TrumpS*_*s*_ or any combination of the three. Hence, the coefficient *δ*_*i*_ identifies the Diff-in-Diff differential effect in a Republican state compared to a Democratic one. Vector *MET*_*st*_ is a set of daily air-quality-station-variant meteorological variables. Weather conditions have been found to be relevant for modelling air pollutant concentrations (*21-25*). As commonly done to capture non-linearities of pressure, precipitation and humidity data, we estimate a quadratic functional form of these variables. Finally, a set of dummy variables that indicates whether a given day is Saturday, Sunday or a bank holiday, similarly to Pearce (*26*), is captured by vector *X*_*t*_. Moreover, we introduce year-month fixed effects to capture longer-run trends that may affect air pollution, *T*_*t*_. A region-quarter fixed effect was also included to account for the heterogeneous geographical distribution of Republican and Democratic states with different climate regimes and seasonal climate fluctuations, *Reg*_*s*_ ∗ *Q*_*t*_. The entire territory is divided into nine zones, the combination of East, Center and West by North, Center and South. These dummy variables are interacted with quarters in order to control for different seasonal effects by territory. We control for this because the political distribution is not uniform throughout the territory and the seasonal effects do not operate at the same time throughout the country (for example, the climatic effect of the beginning and end of spring is not the same in the north and the south), and may have an effect on our estimate. In order not to have an identification problem, we make sure that both parties are represented in all nine zones. This control proves to be very important, as the Covid-19 outbreak took place in late winter and early spring, which is usually a transition period associated with marked changes in the prevailing pollutant levels (*27*). Finally, *α*_*s*_ is the station fixed effect and *ε*_*st*_ is an error term with zero mean, conditional on the monitoring station and the time period. Descriptive statistics of the variables are displayed in Table S1, in the Appendix.

Additionally, we extended the model to include air quality stations fixed effects by post periods P1, P2 and P3, obtaining:

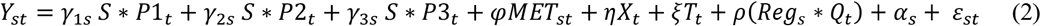

where *S* is the vector of unit-specific dummy variables, *P*1_*t*_, *P*2_*t*_ and *P*3_*t*_ are dummy variables defined as before and *γ*_1*s*_, *γ*_2*s*_ and *γ*_3*s*_ are vectors of station fixed effects associated with these three periods.

The reference model analyses the period from January 1, 2018 to June 30, 2020. This period covers the toughest lockdown period. To check the robustness of the results, we have shortened the analysis period starting one year later (January 1, 2019) and we focused on the same months available for 2020, considering only January to June observations for the periods 2018-2020 and 2019-2020. The interest variables remain statistically significant.

## Supporting information

Supplementary results and Stata code

## Data Availability

Data and Stata code are available under request.

## Appendix

**Table S1.**
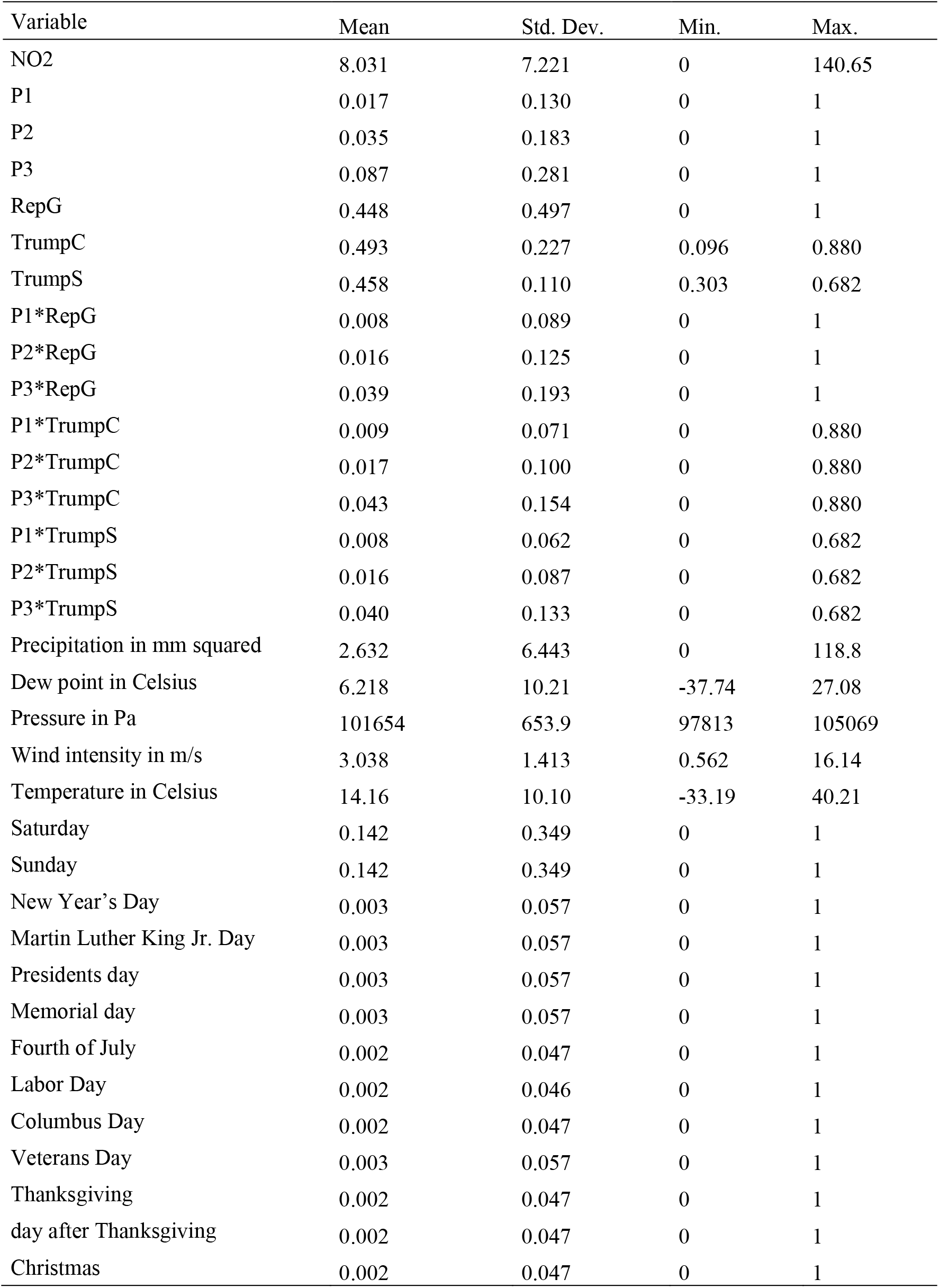
Descriptive statistics.

## Notes

† This research was supported by the grants #ECO2017-86402-C2-1-R and PID2019-104619RB-C42 from the Spanish Ministry of Economy, Industry and Competitiveness.

### Competing Interest Statement

The authors have declared no competing interest.

### Funding Statement

This research was supported by the grants #ECO2017-86402-C2-1-R and PID2019-104619RB-C42 from the Spanish Ministry of Economy, Industry and Competitiveness.

