## Supplementary results and Stata code for "Ideology, policy decision-making and environmental impact in the face of the Coronavirus pandemic in the US"

**This PDF file includes:**

- **Table S1. Descriptive statistics of the whole sample: January 1st 2018 to June 30th 2020**
- **Tables S2 to S9. Different specifications of the Diff-in-Diff models are presented in these tables, basically using different definitions of the sample period and of the post-outbreak periods.**
- **Stata Code**

**Table S1. Descriptive statistics**

| Variable | Mean | Std. Dev. | Min. | Max. |
|---|---|---|---|---|
| NO2 | 8.031 | 7.221 | 0 | 140.65 |
| P1 | 0.017 | 0.130 | 0 | 1 |
| P2 | 0.035 | 0.183 | 0 | 1 |
| P3 | 0.087 | 0.281 | 0 | 1 |
| RepG | 0.448 | 0.497 | 0 | 1 |
| TrumpC | 0.493 | 0.227 | 0.096 | 0.880 |
| TrumpS | 0.458 | 0.110 | 0.303 | 0.682 |
| P1*RepG | 0.008 | 0.089 | 0 | 1 |
| P2*RepG | 0.016 | 0.125 | 0 | 1 |
| P3*RepG | 0.039 | 0.193 | 0 | 1 |
| P1*TrumpC | 0.009 | 0.071 | 0 | 0.880 |
| P2*TrumpC | 0.017 | 0.100 | 0 | 0.880 |
| P3*TrumpC | 0.043 | 0.154 | 0 | 0.880 |
| P1*TrumpS | 0.008 | 0.062 | 0 | 0.682 |
| P2*TrumpS | 0.016 | 0.087 | 0 | 0.682 |
| P3*TrumpS | 0.040 | 0.133 | 0 | 0.682 |
| Precipitation in mm squared | 2.632 | 6.443 | 0 | 118.8 |
| Dew point in Celsius | 6.218 | 10.21 | -37.74 | 27.08 |
| Pressure in Pa | 101654 | 653.9 | 97813 | 105069 |
| Wind intensity in m/s | 3.038 | 1.413 | 0.562 | 16.14 |
| Temperature in Celsius | 14.16 | 10.10 | -33.19 | 40.21 |
| Saturday | 0.142 | 0.349 | 0 | 1 |
| Sunday | 0.142 | 0.349 | 0 | 1 |
| New Year’s Day | 0.003 | 0.057 | 0 | 1 |
| Martin Luther King Jr. Day | 0.003 | 0.057 | 0 | 1 |
| Presidents day | 0.003 | 0.057 | 0 | 1 |
| Memorial day | 0.003 | 0.057 | 0 | 1 |
| Fourth of July | 0.002 | 0.047 | 0 | 1 |
| Labor Day | 0.002 | 0.046 | 0 | 1 |
| Columbus Day | 0.002 | 0.047 | 0 | 1 |
| Veterans Day | 0.003 | 0.057 | 0 | 1 |
| Thanksgiving | 0.002 | 0.047 | 0 | 1 |
| day after Thanksgiving | 0.002 | 0.047 | 0 | 1 |
| Christmas | 0.002 | 0.047 | 0 | 1 |

**Table S2. Station Fixed effect models using data from January 1st, 2018 to June 30th 2020**

| | NO2 | NO2 | NO2 | NO2 | NO2 | NO2 | NO2 |
|---|---|---|---|---|---|---|---|
| P1 | -3.775*** | -3.793*** | -1.821*** | -4.099*** | -1.605*** | -1.945*** | -4.084*** |
| | [-10.00] | [-10.02] | [-8.75] | [-10.64] | [-10.82] | [-9.06] | [-10.71] |
| P2 | -6.487*** | -6.491*** | -3.925*** | -6.577*** | -3.388*** | -4.015*** | -6.579*** |
| | [-14.64] | [-14.72] | [-13.67] | [-15.46] | [-16.10] | [-14.13] | [-15.46] |
| P3 | -4.932*** | -4.945*** | -3.298*** | -4.867*** | -3.066*** | -3.328*** | -4.837*** |
| | [-10.51] | [-10.60] | [-11.82] | [-11.38] | [-14.77] | [-12.16] | [-11.37] |
| P1*RepG | .5175*** | .4308** | .92*** | | 1.061*** | | |
| | [2.87] | [2.59] | [4.54] | | [6.35] | | |
| P2*RepG | .1414 | .1274 | .6525** | | 1.001*** | | |
| | [0.56] | [0.57] | [2.48] | | [4.95] | | |
| P3*RepG | -.1106 | -.1659 | .1949 | | .3422* | | |
| | [-0.44] | [-0.74] | [0.81] | | [1.92] | | |
| P1*TrumpC | -.5847 | | .566 | -.1895 | | 1.653*** | |
| | [-1.36] | | [1.34] | [-0.46] | | [4.65] | |
| P2*TrumpC | -.0936 | | 1.404** | .0153 | | 2.177*** | |
| | [-0.16] | | [2.40] | [0.03] | | [4.87] | |
| P3*TrumpC | -.3637 | | .5996 | -.4539 | | .8331** | |
| | [-0.65] | | [1.09] | [-0.91] | | [2.04] | |
| P1*TrumpS | 5.898*** | 5.393*** | | 6.686*** | | | 6.451*** |
| | [6.30] | [6.58] | | [6.92] | | | [8.28] |
| P2*TrumpS | 7.692*** | 7.612*** | | 7.908*** | | | 7.929*** |
| | [7.84] | [8.31] | | [8.24] | | | [10.29] |
| P3*TrumpS | 4.892*** | 4.584*** | | 4.738*** | | | 4.187*** |
| | [4.71] | [4.61] | | [4.97] | | | [5.59] |

Table S2. (continued)

| | NO2 | NO2 | NO2 | NO2 | NO2 | NO2 | NO2 |
|---|---|---|---|---|---|---|---|
| Precipitation in mm | 2.8e-04 | 2.7e-04 | -2.5e-05 | 3.1e-04 | -1.8e-04 | -7.7e-05 | 2.7e-04 |
| | [0.04] | [0.04] | [-0.00] | [0.04] | [-0.03] | [-0.01] | [0.04] |
| Precipitation in mm ^2 | 2.8e-04*** | 2.8e-04*** | 2.9e-04*** | 2.8e-04*** | 2.9e-04*** | 2.9e-04*** | 2.8e-04*** |
| | [2.77] | [2.77] | [2.82] | [2.76] | [2.84] | [2.82] | [2.77] |
| Dew point in Celsius | -.2187*** | -.2187*** | -.2192*** | -.2186*** | -.2192*** | -.2192*** | -.2185*** |
| | [-15.12] | [-15.13] | [-15.17] | [-15.09] | [-15.20] | [-15.14] | [-15.10] |
| Dew point in Celsius ^2 | -.0025*** | -.0025*** | -.0026*** | -.0025*** | -.0026*** | -.0026*** | -.0025*** |
| | [-6.38] | [-6.39] | [-6.48] | [-6.37] | [-6.45] | [-6.48] | [-6.38] |
| Pressure in Pa | .0222*** | .0222*** | .0219*** | .0221*** | .022*** | .0218*** | .0222*** |
| | [3.29] | [3.30] | [3.23] | [3.28] | [3.25] | [3.22] | [3.29] |
| Pressure in Pa ^2 | -1.1e-07*** | -1.1e-07*** | -1.1e-07*** | -1.1e-07*** | -1.1e-07*** | -1.1e-07*** | -1.1e-07*** |
| | [-3.24] | [-3.25] | [-3.19] | [-3.24] | [-3.21] | [-3.18] | [-3.25] |
| Wind intensity in m/s | -3.05*** | -3.05*** | -3.051*** | -3.051*** | -3.05*** | -3.051*** | -3.051*** |
| | [-23.48] | [-23.47] | [-23.49] | [-23.48] | [-23.48] | [-23.49] | [-23.47] |
| Wind intensity in m/s ^2 | .1953*** | .1952*** | .1953*** | .1953*** | .1952*** | .1954*** | .1953*** |
| | [14.67] | [14.67] | [14.67] | [14.67] | [14.66] | [14.67] | [14.67] |
| Temperature in Celsius | .2082*** | .2081*** | .2095*** | .2081*** | .2096*** | .2098*** | .2078*** |
| | [11.87] | [11.88] | [11.92] | [11.85] | [11.95] | [11.90] | [11.85] |
| Saturday | -1.491*** | -1.491*** | -1.491*** | -1.491*** | -1.491*** | -1.491*** | -1.491*** |
| | [-19.84] | [-19.84] | [-19.84] | [-19.84] | [-19.84] | [-19.84] | [-19.84] |
| Sunday | -2.332*** | -2.332*** | -2.332*** | -2.332*** | -2.332*** | -2.332*** | -2.332*** |
| | [-22.82] | [-22.82] | [-22.82] | [-22.82] | [-22.82] | [-22.82] | [-22.82] |
| New Year's Day | -3.413*** | -3.413*** | -3.41*** | -3.413*** | -3.41*** | -3.41*** | -3.413*** |
| | [-19.42] | [-19.42] | [-19.41] | [-19.42] | [-19.41] | [-19.41] | [-19.42] |
| Martin Luther King Jr. Day | -1.148*** | -1.148*** | -1.145*** | -1.148*** | -1.145*** | -1.145*** | -1.149*** |
| | [-8.39] | [-8.39] | [-8.38] | [-8.39] | [-8.37] | [-8.38] | [-8.39] |
| Presidents day | -1.372*** | -1.372*** | -1.373*** | -1.372*** | -1.373*** | -1.373*** | -1.373*** |
| | [-11.11] | [-11.11] | [-11.12] | [-11.11] | [-11.12] | [-11.12] | [-11.11] |
| Memorial day | -2.464*** | -2.463*** | -2.462*** | -2.463*** | -2.462*** | -2.463*** | -2.463*** |
| | [-17.45] | [-17.45] | [-17.44] | [-17.44] | [-17.44] | [-17.43] | [-17.45] |
| Fourth of July | -1.73*** | -1.73*** | -1.729*** | -1.73*** | -1.729*** | -1.728*** | -1.73*** |
| | [-13.24] | [-13.24] | [-13.23] | [-13.24] | [-13.23] | [-13.23] | [-13.24] |
| Labor Day | -2.882*** | -2.881*** | -2.88*** | -2.882*** | -2.881*** | -2.88*** | -2.881*** |
| | [-17.41] | [-17.41] | [-17.41] | [-17.40] | [-17.42] | [-17.41] | [-17.41] |
| Columbus Day | -1.044*** | -1.044*** | -1.044*** | -1.044*** | -1.045*** | -1.044*** | -1.044*** |
| | [-8.38] | [-8.38] | [-8.38] | [-8.38] | [-8.39] | [-8.38] | [-8.38] |
| Veterans Day | -.4773*** | -.4773*** | -.476*** | -.4776*** | -.4748*** | -.4761*** | -.4776*** |
| | [-3.01] | [-3.01] | [-3.00] | [-3.01] | [-2.99] | [-3.00] | [-3.01] |
| Thanksgiving | -4.266*** | -4.267*** | -4.264*** | -4.266*** | -4.263*** | -4.264*** | -4.267*** |
| | [-20.00] | [-20.00] | [-20.00] | [-20.00] | [-19.99] | [-20.01] | [-20.00] |
| day after Thanksgiving | -2.742*** | -2.743*** | -2.739*** | -2.743*** | -2.739*** | -2.739*** | -2.743*** |
| | [-17.17] | [-17.16] | [-17.17] | [-17.17] | [-17.15] | [-17.17] | [-17.16] |
| Christmas | -4.128*** | -4.129*** | -4.13*** | -4.128*** | -4.13*** | -4.129*** | -4.128*** |
| | [-19.24] | [-19.24] | [-19.24] | [-19.24] | [-19.24] | [-19.24] | [-19.24] |
| N | 311107 | 311107 | 311107 | 311107 | 311107 | 311107 | 311107 |
| R-sq | 0.721 | 0.721 | 0.720 | 0.721 | 0.720 | 0.720 | 0.721 |
| adj. R-sq | 0.720 | 0.720 | 0.720 | 0.720 | 0.720 | 0.720 | 0.720 |
| AIC | 1716309.8 | 1716313.4 | 1716886.0 | 1716324.6 | 1716949.7 | 1716988.2 | 1716329.9 |
| BIC | 1717236.1 | 1717207.8 | 1717780.5 | 1717219.0 | 1717812.2 | 1717850.7 | 1717192.4 |

t statistics in brackets * p<0.10, ** p<0.05, *** p<0.01

**Table S3. Station Fixed effect models using data from January 1st, 2019 to June 30th 2020**

| | NO2 | NO2 | NO2 | NO2 | NO2 | NO2 | NO2 |
|---|---|---|---|---|---|---|---|
| P1 | -3.349*** | -3.379*** | -1.519*** | -3.693*** | -1.498*** | -1.643*** | -3.645*** |
| | [-9.33] | [-9.38] | [-7.58] | [-9.90] | [-10.10] | [-7.93] | [-9.87] |
| P2 | -6.252*** | -6.275*** | -3.667*** | -6.38*** | -3.327*** | -3.763*** | -6.353*** |
| | [-14.55] | [-14.71] | [-12.94] | [-15.74] | [-15.83] | [-13.48] | [-15.66] |
| P3 | -4.715*** | -4.742*** | -3.019*** | -4.697*** | -2.955*** | -3.054*** | -4.643*** |
| | [-9.56] | [-9.67] | [-10.86] | [-10.59] | [-14.27] | [-11.25] | [-10.55] |
| P1*RepG | .5513*** | .3989** | .9286*** | | .9413*** | | |
| | [3.16] | [2.47] | [4.68] | | [5.77] | | |
| P2*RepG | .2031 | .1097 | .7065*** | | .926*** | | |
| | [0.80] | [0.49] | [2.70] | | [4.72] | | |
| P3*RepG | -.0362 | -.161 | .2521 | | .2894* | | |
| | [-0.14] | [-0.71] | [1.05] | | [1.67] | | |
| P1*TrumpC | -1.023** | | .0532 | -.6004 | | 1.15*** | |
| | [-2.39] | | [0.13] | [-1.45] | | [3.33] | |
| P2*TrumpC | -.6219 | | .8846 | -.4644 | | 1.721*** | |
| | [-1.08] | | [1.51] | [-0.91] | | [3.89] | |
| P3*TrumpC | -.8256 | | .1579 | -.8567* | | .4568 | |
| | [-1.47] | | [0.29] | [-1.72] | | [1.12] | |
| P1*TrumpS | 5.524*** | 4.64*** | | 6.359*** | | | 5.613*** |
| | [6.09] | [5.99] | | [6.69] | | | [7.44] |
| P2*TrumpS | 7.743*** | 7.217*** | | 8.051*** | | | 7.493*** |
| | [8.06] | [8.05] | | [8.71] | | | [10.34] |
| P3*TrumpS | 5.033*** | 4.331*** | | 4.991*** | | | 3.958*** |
| | [4.56] | [4.08] | | [4.98] | | | [5.04] |

Table S3. (continued)

| | NO2 | NO2 | NO2 | NO2 | NO2 | NO2 | NO2 |
|---|---|---|---|---|---|---|---|
| Precipitation in mm | -.0103 | -.0103 | -.0109 | -.0103 | -.011 | -.011 | -.0103 |
| | [-1.46] | [-1.46] | [-1.54] | [-1.45] | [-1.56] | [-1.55] | [-1.46] |
| Precipitation in mm ^2 | 3.9e-04*** | 3.9e-04*** | 4.0e-04*** | 3.9e-04*** | 4.0e-04*** | 4.0e-04*** | 3.9e-04*** |
| | [3.77] | [3.77] | [3.83] | [3.76] | [3.85] | [3.83] | [3.77] |
| Dew point in Celsius | -.2014*** | -.2011*** | -.2014*** | -.2013*** | -.2014*** | -.2012*** | -.2008*** |
| | [-13.64] | [-13.68] | [-13.68] | [-13.61] | [-13.75] | [-13.66] | [-13.63] |
| Dew point in Celsius ^2 | -.002*** | -.002*** | -.0021*** | -.002*** | -.0021*** | -.0021*** | -.002*** |
| | [-4.87] | [-4.88] | [-4.99] | [-4.86] | [-4.95] | [-4.99] | [-4.87] |
| Pressure in Pa | .0042 | .0043 | .0035 | .0041 | .0037 | .0033 | .0043 |
| | [0.67] | [0.68] | [0.56] | [0.65] | [0.59] | [0.53] | [0.68] |
| Pressure in Pa ^2 | -1.9e-08 | -2.0e-08 | -1.6e-08 | -1.9e-08 | -1.7e-08 | -1.5e-08 | -2.0e-08 |
| | [-0.63] | [-0.64] | [-0.52] | [-0.61] | [-0.55] | [-0.48] | [-0.64] |
| Wind intensity in m/s | -2.944*** | -2.945*** | -2.948*** | -2.945*** | -2.946*** | -2.948*** | -2.946*** |
| | [-23.88] | [-23.87] | [-23.93] | [-23.87] | [-23.91] | [-23.92] | [-23.87] |
| Wind intensity in m/s ^2 | .1875*** | .1876*** | .1879*** | .1876*** | .1877*** | .188*** | .1877*** |
| | [15.03] | [15.04] | [15.05] | [15.03] | [15.03] | [15.06] | [15.03] |
| Temperature in Celsius | .1882*** | .1874*** | .1898*** | .1881*** | .1898*** | .19*** | .187*** |
| | [10.79] | [10.79] | [10.87] | [10.77] | [10.90] | [10.85] | [10.74] |
| Saturday | -1.432*** | -1.432*** | -1.432*** | -1.432*** | -1.432*** | -1.432*** | -1.432*** |
| | [-18.95] | [-18.95] | [-18.95] | [-18.95] | [-18.95] | [-18.95] | [-18.95] |
| Sunday | -2.24*** | -2.24*** | -2.24*** | -2.24*** | -2.24*** | -2.24*** | -2.24*** |
| | [-22.40] | [-22.40] | [-22.40] | [-22.40] | [-22.40] | [-22.40] | [-22.40] |
| New Year’s Day | -3.614*** | -3.614*** | -3.613*** | -3.614*** | -3.614*** | -3.613*** | -3.614*** |
| | [-19.22] | [-19.22] | [-19.21] | [-19.21] | [-19.22] | [-19.20] | [-19.22] |
| Martin Luther King Jr. Day | -1.475*** | -1.476*** | -1.469*** | -1.475*** | -1.468*** | -1.468*** | -1.476*** |
| | [-8.83] | [-8.83] | [-8.80] | [-8.83] | [-8.80] | [-8.80] | [-8.83] |
| Presidents day | -1.47*** | -1.47*** | -1.471*** | -1.47*** | -1.471*** | -1.47*** | -1.47*** |
| | [-11.49] | [-11.49] | [-11.50] | [-11.49] | [-11.50] | [-11.50] | [-11.49] |
| Memorial day | -2.331*** | -2.33*** | -2.33*** | -2.331*** | -2.33*** | -2.331*** | -2.33*** |
| | [-17.54] | [-17.54] | [-17.52] | [-17.53] | [-17.53] | [-17.51] | [-17.53] |
| Fourth of July | -1.56*** | -1.561*** | -1.559*** | -1.56*** | -1.559*** | -1.558*** | -1.561*** |
| | [-10.41] | [-10.42] | [-10.41] | [-10.41] | [-10.41] | [-10.41] | [-10.42] |
| Labor Day | -2.942*** | -2.94*** | -2.942*** | -2.942*** | -2.942*** | -2.942*** | -2.94*** |
| | [-16.54] | [-16.54] | [-16.54] | [-16.53] | [-16.56] | [-16.54] | [-16.53] |
| Columbus Day | -.3702** | -.3705** | -.3715** | -.37** | -.3721** | -.3717** | -.3703** |
| | [-2.23] | [-2.23] | [-2.24] | [-2.22] | [-2.24] | [-2.24] | [-2.23] |
| Veterans Day | -.3791** | -.3795** | -.3777** | -.3796** | -.3768** | -.3781** | -.38** |
| | [-2.40] | [-2.40] | [-2.39] | [-2.40] | [-2.38] | [-2.39] | [-2.40] |
| Thanksgiving | -4.75*** | -4.751*** | -4.747*** | -4.75*** | -4.746*** | -4.747*** | -4.752*** |
| | [-20.78] | [-20.79] | [-20.78] | [-20.79] | [-20.78] | [-20.79] | [-20.80] |
| day after Thanksgiving | -3.646*** | -3.648*** | -3.641*** | -3.646*** | -3.641*** | -3.641*** | -3.649*** |
| | [-17.82] | [-17.84] | [-17.81] | [-17.82] | [-17.81] | [-17.82] | [-17.83] |
| Christmas | -3.539*** | -3.539*** | -3.541*** | -3.538*** | -3.542*** | -3.541*** | -3.539*** |
| | [-13.79] | [-13.80] | [-13.80] | [-13.79] | [-13.81] | [-13.80] | [-13.80] |
| N | 187138 | 187138 | 187138 | 187138 | 187138 | 187138 | 187138 |
| R-sq | 0.721 | 0.721 | 0.720 | 0.721 | 0.720 | 0.720 | 0.721 |
| adj. R-sq | 0.720 | 0.720 | 0.719 | 0.720 | 0.719 | 0.719 | 0.720 |
| AIC | 1023416.1 | 1023453.7 | 1023951.6 | 1023432.5 | 1023966.4 | 1024064.0 | 1023468.3 |
| BIC | 1024176.5 | 1024183.8 | 1024681.7 | 1024162.5 | 1024666.0 | 1024763.7 | 1024167.9 |

t statistics in brackets * p<0.10, ** p<0.05, *** p<0.01

### Table S4. Station Fixed effect models using data from January to June, 2018 to 2020

| | NO2 | NO2 | NO2 | NO2 | NO2 | NO2 | NO2 |
|---|---|---|---|---|---|---|---|
| P1 | -3.492*** | -3.512*** | -1.659*** | -3.814*** | -1.505*** | -1.784*** | -3.79*** |
| | [-9.06] | [-9.08] | [-7.89] | [-9.71] | [-9.97] | [-8.27] | [-9.77] |
| P2 | -6.324*** | -6.333*** | -3.823*** | -6.412*** | -3.356*** | -3.916*** | -6.402*** |
| | [-14.19] | [-14.29] | [-13.15] | [-14.96] | [-15.73] | [-13.69] | [-14.97] |
| P3 | -4.745*** | -4.762*** | -3.199*** | -4.687*** | -3.022*** | -3.234*** | -4.65*** |
| | [-10.02] | [-10.12] | [-11.16] | [-10.86] | [-14.30] | [-11.54] | [-10.84] |
| P1*RepG | .5144*** | .4124** | .9156*** | | 1.019*** | | |
| | [2.70] | [2.35] | [4.30] | | [5.95] | | |
| P2*RepG | .1369 | .0989 | .6612** | | .9681*** | | |
| | [0.52] | [0.44] | [2.42] | | [4.76] | | |
| P3*RepG | -.1012 | -.1711 | .2127 | | .3288* | | |
| | [-0.38] | [-0.72] | [0.84] | | [1.78] | | |
| P1*TrumpC | -.6846 | | .4059 | -.2926 | | 1.492*** | |
| | [-1.52] | | [0.93] | [-0.68] | | [4.16] | |
| P2*TrumpC | -.2527 | | 1.223** | -.148 | | 2.012*** | |
| | [-0.43] | | [2.04] | [-0.29] | | [4.53] | |
| P3*TrumpC | -.4608 | | .4619 | -.5437 | | .7216* | |
| | [-0.79] | | [0.81] | [-1.06] | | [1.73] | |
| P1*TrumpS | 5.568*** | 4.978*** | | 6.354*** | | | 5.989*** |
| | [5.77] | [5.95] | | [6.36] | | | [7.62] |
| P2*TrumpS | 7.547*** | 7.333*** | | 7.759*** | | | 7.58*** |
| | [7.67] | [8.01] | | [7.96] | | | [9.93] |
| P3*TrumpS | 4.664*** | 4.275*** | | 4.525*** | | | 3.865*** |
| | [4.47] | [4.28] | | [4.70] | | | [5.20] |
| Precipitation in mm | -2.9e-04 | -4.0e-04 | -8.8e-04 | -2.6e-04 | -9.5e-04 | -.0011 | -4.1e-04 |
| | [-0.04] | [-0.05] | [-0.11] | [-0.03] | [-0.12] | [-0.14] | [-0.05] |
| Precipitation in mm ^2 | 2.7e-04** | 2.7e-04** | 2.8e-04** | 2.7e-04** | 2.8e-04** | 2.8e-04** | 2.7e-04** |
| | [2.18] | [2.18] | [2.23] | [2.17] | [2.23] | [2.26] | [2.18] |
| Dew point in Celsius | -.1943*** | -.1941*** | -.1944*** | -.1941*** | -.1944*** | -.1943*** | -.1939*** |
| | [-12.78] | [-12.80] | [-12.81] | [-12.75] | [-12.86] | [-12.79] | [-12.77] |
| Dew point in Celsius ^2 | -.0021*** | -.0021*** | -.0022*** | -.0021*** | -.0022*** | -.0022*** | -.0021*** |
| | [-5.09] | [-5.10] | [-5.25] | [-5.08] | [-5.22] | [-5.27] | [-5.09] |
| Pressure in Pa | .017*** | .017*** | .0166** | .017*** | .0169** | .0165** | .017*** |
| | [2.61] | [2.62] | [2.54] | [2.60] | [2.58] | [2.52] | [2.61] |
| Pressure in Pa ^2 | -8.2e-08** | -8.2e-08** | -8.0e-08** | -8.2e-08** | -8.2e-08** | -8.0e-08** | -8.2e-08** |
| | [-2.57] | [-2.57] | [-2.50] | [-2.56] | [-2.53] | [-2.48] | [-2.57] |
| Wind intensity in m/s | -2.984*** | -2.985*** | -2.987*** | -2.985*** | -2.985*** | -2.988*** | -2.986*** |
| | [-23.53] | [-23.50] | [-23.56] | [-23.53] | [-23.53] | [-23.56] | [-23.50] |
| Wind intensity in m/s ^2 | .1874*** | .1874*** | .1877*** | .1875*** | .1875*** | .1878*** | .1875*** |
| | [14.81] | [14.79] | [14.83] | [14.81] | [14.79] | [14.83] | [14.79] |
| Temperature in Celsius | .1958*** | .1954*** | .197*** | .1956*** | .1972*** | .1973*** | .195*** |
| | [11.09] | [11.11] | [11.14] | [11.06] | [11.18] | [11.12] | [11.07] |
| Saturday | -1.498*** | -1.498*** | -1.498*** | -1.498*** | -1.498*** | -1.498*** | -1.498*** |
| | [-19.27] | [-19.27] | [-19.27] | [-19.27] | [-19.27] | [-19.27] | [-19.27] |
| Sunday | -2.339*** | -2.339*** | -2.338*** | -2.339*** | -2.338*** | -2.338*** | -2.339*** |
| | [-22.57] | [-22.57] | [-22.57] | [-22.57] | [-22.57] | [-22.58] | [-22.58] |
| New Year's Day | -3.401*** | -3.402*** | -3.396*** | -3.401*** | -3.396*** | -3.396*** | -3.402*** |
| | [-19.53] | [-19.52] | [-19.53] | [-19.53] | [-19.50] | [-19.52] | [-19.52] |
| Martin Luther King Jr. Day | -1.119*** | -1.119*** | -1.114*** | -1.119*** | -1.113*** | -1.114*** | -1.12*** |
| | [-8.41] | [-8.41] | [-8.40] | [-8.42] | [-8.38] | [-8.40] | [-8.42] |
| Presidents day | -1.395*** | -1.395*** | -1.396*** | -1.395*** | -1.396*** | -1.396*** | -1.395*** |
| | [-11.30] | [-11.30] | [-11.31] | [-11.30] | [-11.31] | [-11.31] | [-11.30] |
| Memorial day | -2.518*** | -2.518*** | -2.516*** | -2.518*** | -2.516*** | -2.516*** | -2.518*** |
| | [-17.66] | [-17.66] | [-17.64] | [-17.65] | [-17.64] | [-17.63] | [-17.66] |
| N | 185671 | 185671 | 185671 | 185671 | 185671 | 185671 | 185671 |
| R-sq | 0.717 | 0.717 | 0.716 | 0.717 | 0.716 | 0.716 | 0.717 |
| adj. R-sq | 0.716 | 0.716 | 0.716 | 0.716 | 0.716 | 0.716 | 0.716 |
| AIC | 1022543.5 | 1022551.8 | 1023046.0 | 1022557.6 | 1023086.6 | 1023144.9 | 1022567.2 |
| BIC | 1023070.3 | 1023048.3 | 1023542.4 | 1023054.0 | 1023552.6 | 1023611.0 | 1023033.2 |

t statistics in brackets * p<0.10, ** p<0.05, *** p<0.01

Note: Holidays of the second half of the year are out of sample and their dummy variables removed

### Table S5. Station Fixed effect models using data from January to June, 2019 to 2020

| | NO2 | NO2 | NO2 | NO2 | NO2 | NO2 | NO2 |
|---|---|---|---|---|---|---|---|
| P1 | -3.047*** | -3.072*** | -1.408*** | -3.394*** | -1.399*** | -1.534*** | -3.347*** |
| | [-8.39] | [-8.41] | [-6.92] | [-8.94] | [-9.29] | [-7.36] | [-8.96] |
| P2 | -6.119*** | -6.143*** | -3.62*** | -6.253*** | -3.299*** | -3.722*** | -6.223*** |
| | [-14.40] | [-14.58] | [-12.72] | [-15.51] | [-15.58] | [-13.35] | [-15.50] |
| P3 | -4.523*** | -4.547*** | -2.973*** | -4.519*** | -2.901*** | -3.016*** | -4.468*** |
| | [-9.16] | [-9.27] | [-10.30] | [-10.19] | [-13.74] | [-10.74] | [-10.15] |
| P1*RepG | .5533*** | .4108** | .9216*** | | .9298*** | | |
| | [3.01] | [2.43] | [4.42] | | [5.61] | | |
| P2*RepG | .2075 | .1122 | .7285*** | | .9383*** | | |
| | [0.82] | [0.51] | [2.74] | | [4.86] | | |
| P3*RepG | -.0189 | -.1288 | .2783 | | .3235* | | |
| | [-0.07] | [-0.55] | [1.11] | | [1.81] | | |
| P1*TrumpC | -.9516** | | .0247 | -.5301 | | 1.119*** | |
| | [-2.06] | | [0.06] | [-1.20] | | [3.20] | |
| P2*TrumpC | -.6367 | | .8387 | -.4781 | | 1.707*** | |
| | [-1.09] | | [1.42] | [-0.94] | | [3.96] | |
| P3*TrumpC | -.7335 | | .1832 | -.7527 | | .5204 | |
| | [-1.26] | | [0.32] | [-1.48] | | [1.26] | |
| P1*TrumpS | 4.988*** | 4.164*** | | 5.834*** | | | 5.168*** |
| | [5.28] | [5.31] | | [5.83] | | | [6.84] |
| P2*TrumpS | 7.536*** | 7.000*** | | 7.859*** | | | 7.282*** |
| | [7.96] | [8.03] | | [8.45] | | | [10.39] |
| P3*TrumpS | 4.651*** | 4.029*** | | 4.643*** | | | 3.734*** |
| | [4.30] | [3.84] | | [4.70] | | | [4.85] |
| Precipitation in mm | -.0092 | -.0094 | -.0099 | -.0092 | -.01 | -.0102 | -.0095 |
| | [-1.20] | [-1.22] | [-1.29] | [-1.20] | [-1.30] | [-1.33] | [-1.23] |
| Precipitation in mm ^2 | 3.9e-04*** | 4.0e-04*** | 4.0e-04*** | 3.9e-04*** | 4.0e-04*** | 4.0e-04*** | 4.0e-04*** |
| | [3.12] | [3.13] | [3.16] | [3.11] | [3.15] | [3.19] | [3.13] |
| Dew point in Celsius | -.1756*** | -.1749*** | -.1749*** | -.1754*** | -.1751*** | -.1746*** | -.1745*** |
| | [-11.16] | [-11.23] | [-11.18] | [-11.12] | [-11.30] | [-11.18] | [-11.16] |
| Dew point in Celsius ^2 | -.0017*** | -.0017*** | -.0018*** | -.0017*** | -.0018*** | -.0018*** | -.0017*** |
| | [-4.06] | [-4.08] | [-4.23] | [-4.05] | [-4.18] | [-4.25] | [-4.07] |
| Pressure in Pa | .0051 | .0052 | .0044 | .005 | .0047 | .0041 | .0051 |
| | [0.88] | [0.89] | [0.76] | [0.86] | [0.79] | [0.70] | [0.88] |
| Pressure in Pa ^2 | -2.4e-08 | -2.4e-08 | -2.1e-08 | -2.3e-08 | -2.2e-08 | -1.9e-08 | -2.4e-08 |
| | [-0.84] | [-0.85] | [-0.72] | [-0.82] | [-0.75] | [-0.66] | [-0.84] |
| Wind intensity in m/s | -2.869*** | -2.871*** | -2.878*** | -2.87*** | -2.876*** | -2.878*** | -2.872*** |
| | [-23.99] | [-23.95] | [-24.09] | [-23.99] | [-24.04] | [-24.09] | [-23.96] |
| Wind intensity in m/s ^2 | .1803*** | .1806*** | .1813*** | .1804*** | .181*** | .1813*** | .1807*** |
| | [15.25] | [15.25] | [15.31] | [15.26] | [15.26] | [15.32] | [15.25] |
| Temperature in Celsius | .1688*** | .1674*** | .1702*** | .1686*** | .1704*** | .1705*** | .1669*** |
| | [9.69] | [9.70] | [9.78] | [9.67] | [9.84] | [9.78] | [9.63] |
| Saturday | -1.407*** | -1.407*** | -1.407*** | -1.407*** | -1.407*** | -1.407*** | -1.407*** |
| | [-18.47] | [-18.47] | [-18.47] | [-18.47] | [-18.47] | [-18.47] | [-18.47] |
| Sunday | -2.212*** | -2.212*** | -2.212*** | -2.212*** | -2.212*** | -2.212*** | -2.212*** |
| | [-22.45] | [-22.45] | [-22.45] | [-22.45] | [-22.45] | [-22.45] | [-22.45] |
| New Year's Day | -3.604*** | -3.604*** | -3.601*** | -3.604*** | -3.602*** | -3.601*** | -3.603*** |
| | [-19.20] | [-19.21] | [-19.19] | [-19.20] | [-19.20] | [-19.19] | [-19.21] |
| Martin Luther King Jr. Day | -1.416*** | -1.418*** | -1.407*** | -1.417*** | -1.406*** | -1.406*** | -1.419*** |
| | [-8.70] | [-8.71] | [-8.65] | [-8.70] | [-8.64] | [-8.66] | [-8.71] |
| Presidents day | -1.466*** | -1.467*** | -1.468*** | -1.466*** | -1.468*** | -1.468*** | -1.467*** |
| | [-11.55] | [-11.54] | [-11.56] | [-11.55] | [-11.56] | [-11.56] | [-11.54] |
| Memorial day | -2.354*** | -2.353*** | -2.353*** | -2.354*** | -2.353*** | -2.355*** | -2.353*** |
| | [-17.77] | [-17.77] | [-17.75] | [-17.76] | [-17.75] | [-17.74] | [-17.76] |
| N | 124418 | 124418 | 124418 | 124418 | 124418 | 124418 | 124418 |
| R-sq | 0.715 | 0.715 | 0.714 | 0.715 | 0.714 | 0.714 | 0.715 |
| adj. R-sq | 0.714 | 0.714 | 0.713 | 0.714 | 0.713 | 0.713 | 0.714 |
| AIC | 675913.8 | 675941.6 | 676359.6 | 675930.2 | 676372.3 | 676470.4 | 675954.6 |
| BIC | 676361.5 | 676360.1 | 676778.1 | 676348.7 | 676761.6 | 676859.6 | 676343.9 |

t statistics in brackets * p<0.10, ** p<0.05, *** p<0.01

Note: Holidays of the second half of the year are out of sample and their dummy variables removed

**Table S6. Station Fixed effect models using data from January 1st, 2018 to June 30th 2020 with different post period specifications**

| | P1 & P2 | | | P2 & P3 | | |
|---|---|---|---|---|---|---|
| | NO2 | NO2 | NO2 | NO2 | NO2 | NO2 |
| P1 | -2.254*** | -2.271*** | -2.584*** | | | |
| | [-6.00] | [-6.04] | [-6.87] | | | |
| P2 | -4.014*** | -4.017*** | -4.107*** | -5.326*** | -5.328*** | -5.408*** |
| | [-10.85] | [-10.92] | [-11.69] | [-13.09] | [-13.16] | [-14.11] |
| P3 | | | | -3.817*** | -3.829*** | -3.716*** |
| | | | | [-8.53] | [-8.62] | [-9.34] |
| P1*RepG | .5428*** | .4576*** | | | | |
| | [3.07] | [2.76] | | | | |
| P2*RepG | .1424 | .1343 | | .1276 | .1151 | |
| | [0.61] | [0.65] | | [0.51] | [0.52] | |
| P3*RepG | | | | -.1171 | -.1707 | |
| | | | | [-0.47] | [-0.76] | |
| P1*TrumpC | -.5761 | | | | | |
| | [-1.38] | | | | | |
| P2*TrumpC | -.0545 | | | -.0831 | | |
| | [-0.10] | | | [-0.15] | | |
| P3*TrumpC | | | | -.3527 | | |
| | | | | [-0.63] | | |
| P1*TrumpS | 5.787*** | 5.289*** | 6.418*** | | | |
| | [6.13] | [6.38] | [8.26] | | | |
| P2*TrumpS | 7.091*** | 7.045*** | 7.374*** | 7.535*** | 7.464*** | 7.752*** |
| | [7.84] | [8.33] | [10.22] | [7.73] | [8.20] | [10.19] |
| P3*TrumpS | | | | 4.834*** | 4.535*** | 4.123*** |
| | | | | [4.65] | [4.56] | [5.51] |

Table S6. (continued)

| | NO2 | NO2 | NO2 | NO2 | NO2 | NO2 |
|---|---|---|---|---|---|---|
| Precipitation in mm | 9.7e-05 | 1.2e-04 | 1.1e-04 | -1.8e-04 | -2.1e-04 | -2.1e-04 |
| | [0.01] | [0.02] | [0.02] | [-0.03] | [-0.03] | [-0.03] |
| Precipitation in mm ^2 | 2.9e-04*** | 2.8e-04*** | 2.8e-04*** | 2.9e-04*** | 2.9e-04*** | 2.9e-04*** |
| | [2.81] | [2.81] | [2.81] | [2.84] | [2.85] | [2.84] |
| Dew point in Celsius | -.2183*** | -.2183*** | -.2183*** | -.217*** | -.2169*** | -.2169*** |
| | [-15.10] | [-15.09] | [-15.09] | [-15.11] | [-15.12] | [-15.09] |
| Dew point in Celsius ^2 | -.0026*** | -.0026*** | -.0026*** | -.0026*** | -.0026*** | -.0026*** |
| | [-6.47] | [-6.49] | [-6.48] | [-6.43] | [-6.44] | [-6.43] |
| Pressure in Pa | .0213*** | .0213*** | .0213*** | .022*** | .022*** | .0221*** |
| | [3.15] | [3.16] | [3.15] | [3.27] | [3.28] | [3.28] |
| Pressure in Pa ^2 | -1.0e-07*** | -1.0e-07*** | -1.0e-07*** | -1.1e-07*** | -1.1e-07*** | -1.1e-07*** |
| | [-3.10] | [-3.11] | [-3.11] | [-3.22] | [-3.23] | [-3.23] |
| Wind intensity in m/s | -3.054*** | -3.054*** | -3.054*** | -3.047*** | -3.047*** | -3.047*** |
| | [-23.50] | [-23.49] | [-23.49] | [-23.45] | [-23.45] | [-23.45] |
| Wind intensity in m/s ^2 | .1956*** | .1956*** | .1956*** | .1949*** | .1949*** | .1949*** |
| | [14.67] | [14.67] | [14.67] | [14.62] | [14.62] | [14.62] |
| Temperature in Celsius | .2068*** | .2068*** | .2067*** | .2067*** | .2065*** | .2064*** |
| | [11.83] | [11.84] | [11.83] | [11.84] | [11.85] | [11.83] |
| Saturday | -1.495*** | -1.495*** | -1.495*** | -1.493*** | -1.493*** | -1.493*** |
| | [-19.88] | [-19.88] | [-19.88] | [-19.85] | [-19.85] | [-19.85] |
| Sunday | -2.323*** | -2.323*** | -2.323*** | -2.327*** | -2.327*** | -2.327*** |
| | [-22.76] | [-22.76] | [-22.76] | [-22.80] | [-22.80] | [-22.80] |
| New Year’s Day | -3.414*** | -3.414*** | -3.414*** | -3.41*** | -3.41*** | -3.41*** |
| | [-19.43] | [-19.43] | [-19.43] | [-19.41] | [-19.41] | [-19.41] |
| Martin Luther King Jr. Day | -1.147*** | -1.147*** | -1.147*** | -1.146*** | -1.146*** | -1.146*** |
| | [-8.38] | [-8.38] | [-8.38] | [-8.37] | [-8.37] | [-8.37] |
| Presidents day | -1.302*** | -1.302*** | -1.302*** | -1.32*** | -1.32*** | -1.32*** |
| | [-10.51] | [-10.51] | [-10.51] | [-10.64] | [-10.65] | [-10.65] |
| Memorial day | -2.46*** | -2.46*** | -2.46*** | -2.463*** | -2.463*** | -2.463*** |
| | [-17.43] | [-17.43] | [-17.43] | [-17.45] | [-17.45] | [-17.44] |
| Fourth of July | -1.73*** | -1.73*** | -1.73*** | -1.73*** | -1.73*** | -1.73*** |
| | [-13.24] | [-13.25] | [-13.25] | [-13.24] | [-13.24] | [-13.25] |
| Labor Day | -2.874*** | -2.874*** | -2.874*** | -2.879*** | -2.879*** | -2.879*** |
| | [-17.39] | [-17.39] | [-17.38] | [-17.40] | [-17.41] | [-17.40] |
| Columbus Day | -1.043*** | -1.043*** | -1.043*** | -1.044*** | -1.044*** | -1.044*** |
| | [-8.37] | [-8.37] | [-8.37] | [-8.37] | [-8.37] | [-8.37] |
| Veterans Day | -.4721*** | -.4721*** | -.4723*** | -.4753*** | -.4754*** | -.4754*** |
| | [-2.97] | [-2.97] | [-2.97] | [-3.00] | [-3.00] | [-3.00] |
| Thanksgiving | -4.269*** | -4.269*** | -4.269*** | -4.268*** | -4.268*** | -4.268*** |
| | [-20.00] | [-20.00] | [-20.00] | [-19.99] | [-19.99] | [-19.99] |
| day after Thanksgiving | -2.745*** | -2.745*** | -2.745*** | -2.746*** | -2.746*** | -2.747*** |
| | [-17.16] | [-17.17] | [-17.17] | [-17.18] | [-17.18] | [-17.17] |
| Christmas | -4.131*** | -4.131*** | -4.131*** | -4.129*** | -4.129*** | -4.13*** |
| | [-19.24] | [-19.24] | [-19.24] | [-19.23] | [-19.23] | [-19.23] |
| N | 311107 | 311107 | 311107 | 311107 | 311107 | 311107 |
| R-sq | 0.720 | 0.720 | 0.720 | 0.721 | 0.720 | 0.720 |
| adj. R-sq | 0.720 | 0.720 | 0.720 | 0.720 | 0.720 | 0.720 |
| AIC | 1716981.3 | 1716980.9 | 1716992.1 | 1716593.6 | 1716595.4 | 1716601.8 |
| BIC | 1717865.0 | 1717843.4 | 1717833.3 | 1717477.4 | 1717457.9 | 1717443.0 |

t statistics in brackets * p<0.10, ** p<0.05, *** p<0.01

**Table S7. Station Fixed effect models using data from January 1st, 2019 to June 30th 2020 with different post period specifications**

| | P1 & P2 | | | P2 & P3 | | |
|---|---|---|---|---|---|---|
| | NO2 | NO2 | NO2 | NO2 | NO2 | NO2 |
| P1 | -1.798*** | -1.826*** | -2.124*** | | | |
| | [-4.89] | [-4.96] | [-5.71] | | | |
| P2 | -3.733*** | -3.751*** | -3.836*** | -5.112*** | -5.133*** | -5.198*** |
| | [-10.97] | [-11.16] | [-11.95] | [-12.95] | [-13.14] | [-14.35] |
| P3 | | | | -3.63*** | -3.655*** | -3.548*** |
| | | | | [-7.65] | [-7.77] | [-8.56] |
| P1*RepG | .5783*** | .4392*** | | | | |
| | [3.41] | [2.72] | | | | |
| P2*RepG | .1992 | .1268 | | .1785 | .0912 | |
| | [0.91] | [0.66] | | [0.72] | [0.41] | |
| P3*RepG | | | | -.0506 | -.1697 | |
| | | | | [-0.20] | [-0.75] | |
| P1*TrumpC | -.941** | | | | | |
| | [-2.28] | | | | | |
| P2*TrumpC | -.4823 | | | -.5809 | | |
| | [-0.97] | | | [-1.02] | | |
| P3*TrumpC | | | | -.7877 | | |
| | | | | [-1.41] | | |
| P1*TrumpS | 5.171*** | 4.357*** | 5.437*** | | | |
| | [5.54] | [5.42] | [7.13] | | | |
| P2*TrumpS | 6.791*** | 6.383*** | 6.692*** | 7.501*** | 7.008*** | 7.24*** |
| | [8.17] | [8.24] | [10.25] | [7.86] | [7.88] | [10.16] |
| P3*TrumpS | | | | 4.912*** | 4.241*** | 3.843*** |
| | | | | [4.45] | [3.99] | [4.88] |

Table S7. (continued)

| | NO2 | NO2 | NO2 | NO2 | NO2 | NO2 |
|---|---|---|---|---|---|---|
| Precipitation in mm | -.0109 | -.0108 | -.0108 | -.011 | -.0111 | -.0111 |
| | [-1.54] | [-1.52] | [-1.52] | [-1.56] | [-1.56] | [-1.56] |
| Precipitation in mm ^2 | 4.0e-04*** | 4.0e-04*** | 4.0e-04*** | 4.0e-04*** | 4.0e-04*** | 4.0e-04*** |
| | [3.85] | [3.85] | [3.85] | [3.87] | [3.88] | [3.88] |
| Dew point in Celsius | -.2002*** | -.2002*** | -.2001*** | -.1984*** | -.1981*** | -.198*** |
| | [-13.62] | [-13.62] | [-13.62] | [-13.62] | [-13.66] | [-13.60] |
| Dew point in Celsius ^2 | -.0021*** | -.0021*** | -.0021*** | -.0021*** | -.0021*** | -.0021*** |
| | [-4.96] | [-5.01] | [-5.00] | [-4.97] | [-4.97] | [-4.96] |
| Pressure in Pa | .0031 | .003 | .0029 | .0038 | .0039 | .0039 |
| | [0.48] | [0.47] | [0.46] | [0.61] | [0.62] | [0.63] |
| Pressure in Pa ^2 | -1.4e-08 | -1.4e-08 | -1.3e-08 | -1.8e-08 | -1.8e-08 | -1.8e-08 |
| | [-0.44] | [-0.43] | [-0.42] | [-0.57] | [-0.58] | [-0.59] |
| Wind intensity in m/s | -2.953*** | -2.953*** | -2.954*** | -2.937*** | -2.938*** | -2.939*** |
| | [-23.92] | [-23.91] | [-23.90] | [-23.84] | [-23.83] | [-23.83] |
| Wind intensity in m/s ^2 | .1883*** | .1884*** | .1884*** | .1868*** | .187*** | .187*** |
| | [15.05] | [15.04] | [15.04] | [14.95] | [14.95] | [14.95] |
| Temperature in Celsius | .1854*** | .1855*** | .1854*** | .1855*** | .1848*** | .1846*** |
| | [10.71] | [10.72] | [10.71] | [10.72] | [10.73] | [10.68] |
| Saturday | -1.439*** | -1.439*** | -1.439*** | -1.436*** | -1.436*** | -1.436*** |
| | [-19.01] | [-19.01] | [-19.01] | [-18.96] | [-18.96] | [-18.96] |
| Sunday | -2.225*** | -2.225*** | -2.225*** | -2.231*** | -2.231*** | -2.231*** |
| | [-22.30] | [-22.29] | [-22.30] | [-22.36] | [-22.37] | [-22.37] |
| New Year’s Day | -3.619*** | -3.619*** | -3.619*** | -3.612*** | -3.611*** | -3.611*** |
| | [-19.24] | [-19.25] | [-19.25] | [-19.23] | [-19.23] | [-19.23] |
| Martin Luther King Jr. Day | -1.473*** | -1.473*** | -1.473*** | -1.47*** | -1.471*** | -1.471*** |
| | [-8.80] | [-8.80] | [-8.80] | [-8.81] | [-8.81] | [-8.82] |
| Presidents day | -1.367*** | -1.367*** | -1.366*** | -1.395*** | -1.395*** | -1.396*** |
| | [-10.75] | [-10.76] | [-10.76] | [-10.87] | [-10.87] | [-10.87] |
| Memorial day | -2.326*** | -2.326*** | -2.326*** | -2.33*** | -2.33*** | -2.329*** |
| | [-17.52] | [-17.52] | [-17.52] | [-17.54] | [-17.54] | [-17.53] |
| Fourth of July | -1.565*** | -1.565*** | -1.565*** | -1.561*** | -1.562*** | -1.562*** |
| | [-10.44] | [-10.44] | [-10.44] | [-10.42] | [-10.42] | [-10.42] |
| Labor Day | -2.93*** | -2.929*** | -2.929*** | -2.935*** | -2.934*** | -2.934*** |
| | [-16.50] | [-16.50] | [-16.50] | [-16.53] | [-16.53] | [-16.52] |
| Columbus Day | -.3754** | -.3751** | -.375** | -.368** | -.3684** | -.3683** |
| | [-2.26] | [-2.25] | [-2.25] | [-2.21] | [-2.21] | [-2.21] |
| Veterans Day | -.3736** | -.3739** | -.3743** | -.3783** | -.3787** | -.3789** |
| | [-2.36] | [-2.36] | [-2.37] | [-2.39] | [-2.39] | [-2.39] |
| Thanksgiving | -4.764*** | -4.764*** | -4.765*** | -4.757*** | -4.758*** | -4.758*** |
| | [-20.81] | [-20.82] | [-20.82] | [-20.79] | [-20.79] | [-20.79] |
| day after Thanksgiving | -3.657*** | -3.657*** | -3.658*** | -3.656*** | -3.658*** | -3.658*** |
| | [-17.87] | [-17.87] | [-17.88] | [-17.87] | [-17.87] | [-17.86] |
| Christmas | -3.548*** | -3.548*** | -3.548*** | -3.54*** | -3.541*** | -3.541*** |
| | [-13.81] | [-13.82] | [-13.82] | [-13.79] | [-13.79] | [-13.79] |
| N | 187138 | 187138 | 187138 | 187138 | 187138 | 187138 |
| R-sq | 0.720 | 0.720 | 0.720 | 0.721 | 0.720 | 0.720 |
| adj. R-sq | 0.719 | 0.719 | 0.719 | 0.720 | 0.720 | 0.720 |
| AIC | 1024042.0 | 1024052.3 | 1024062.6 | 1023662.1 | 1023689.6 | 1023695.2 |
| BIC | 1024761.9 | 1024752.0 | 1024741.9 | 1024382.0 | 1024389.3 | 1024374.5 |

t statistics in brackets * p<0.10, ** p<0.05, *** p<0.01

**Table S8. Station Fixed effect models using data from January to June, 2018 to 2020 with different post period specifications**

| | P1 & P2 | | | P2 & P3 | | |
|---|---|---|---|---|---|---|
| | NO2 | NO2 | NO2 | NO2 | NO2 | NO2 |
| P1 | -1.929*** | -1.948*** | -2.26*** | | | |
| | [-5.00] | [-5.04] | [-5.92] | | | |
| P2 | -3.839*** | -3.846*** | -3.931*** | -5.235*** | -5.244*** | -5.301*** |
| | [-10.70] | [-10.80] | [-11.46] | [-12.87] | [-12.97] | [-13.85] |
| P3 | | | | -3.695*** | -3.71*** | -3.59*** |
| | | | | [-8.23] | [-8.33] | [-9.02] |
| P1*RepG | .5513*** | .4558*** | | | | |
| | [3.01] | [2.64] | | | | |
| P2*RepG | .1521 | .1244 | | .1183 | .0832 | |
| | [0.67] | [0.63] | | [0.46] | [0.37] | |
| P3*RepG | | | | -.1128 | -.179 | |
| | | | | [-0.43] | [-0.76] | |
| P1*TrumpC | -.6458 | | | | | |
| | [-1.50] | | | | | |
| P2*TrumpC | -.1845 | | | -.233 | | |
| | [-0.36] | | | [-0.40] | | |
| P3*TrumpC | | | | -.4369 | | |
| | | | | [-0.76] | | |
| P1*TrumpS | 5.279*** | 4.721*** | 5.848*** | | | |
| | [5.37] | [5.51] | [7.45] | | | |
| P2*TrumpS | 6.806*** | 6.65*** | 6.957*** | 7.368*** | 7.171*** | 7.378*** |
| | [7.80] | [8.17] | [9.90] | [7.56] | [7.91] | [9.82] |
| P3*TrumpS | | | | 4.566*** | 4.197*** | 3.762*** |
| | | | | [4.38] | [4.20] | [5.06] |
| Precipitation in mm | -8.5e-04 | -8.2e-04 | -8.3e-04 | -.0012 | -.0013 | -.0013 |
| | [-0.11] | [-0.11] | [-0.11] | [-0.15] | [-0.17] | [-0.17] |
| Precipitation in mm ^2 | 2.9e-04** | 2.8e-04** | 2.9e-04** | 2.9e-04** | 2.9e-04** | 2.9e-04** |
| | [2.26] | [2.26] | [2.27] | [2.31] | [2.31] | [2.31] |
| Dew point in Celsius | -.193*** | -.1931*** | -.193*** | -.1913*** | -.1912*** | -.191*** |
| | [-12.74] | [-12.73] | [-12.73] | [-12.76] | [-12.78] | [-12.75] |
| Dew point in Celsius ^2 | -.0022*** | -.0022*** | -.0022*** | -.0021*** | -.0021*** | -.0021*** |
| | [-5.27] | [-5.29] | [-5.28] | [-5.17] | [-5.18] | [-5.17] |
| Pressure in Pa | .0157** | .0157** | .0157** | .0168** | .0168** | .0168** |
| | [2.39] | [2.40] | [2.39] | [2.57] | [2.58] | [2.58] |
| Pressure in Pa ^2 | -7.6e-08** | -7.6e-08** | -7.6e-08** | -8.1e-08** | -8.1e-08** | -8.1e-08** |
| | [-2.35] | [-2.36] | [-2.35] | [-2.53] | [-2.53] | [-2.54] |
| Wind intensity in m/s | -2.99*** | -2.99*** | -2.991*** | -2.979*** | -2.979*** | -2.98*** |
| | [-23.55] | [-23.53] | [-23.53] | [-23.48] | [-23.45] | [-23.46] |
| Wind intensity in m/s ^2 | .1879*** | .1879*** | .1879*** | .1868*** | .1869*** | .1869*** |
| | [14.81] | [14.79] | [14.80] | [14.73] | [14.71] | [14.71] |
| Temperature in Celsius | .1929*** | .193*** | .1928*** | .1931*** | .1928*** | .1926*** |
| | [10.98] | [10.99] | [10.99] | [11.04] | [11.06] | [11.03] |
| Saturday | -1.504*** | -1.504*** | -1.504*** | -1.502*** | -1.501*** | -1.501*** |
| | [-19.33] | [-19.33] | [-19.33] | [-19.28] | [-19.28] | [-19.28] |
| Sunday | -2.323*** | -2.323*** | -2.323*** | -2.331*** | -2.331*** | -2.331*** |
| | [-22.48] | [-22.48] | [-22.48] | [-22.54] | [-22.55] | [-22.55] |
| New Year's Day | -3.402*** | -3.402*** | -3.402*** | -3.397*** | -3.398*** | -3.398*** |
| | [-19.54] | [-19.55] | [-19.55] | [-19.53] | [-19.51] | [-19.51] |
| Martin Luther King Jr. Day | -1.117*** | -1.117*** | -1.117*** | -1.116*** | -1.117*** | -1.117*** |
| | [-8.39] | [-8.39] | [-8.39] | [-8.40] | [-8.39] | [-8.40] |
| Presidents day | -1.326*** | -1.326*** | -1.326*** | -1.347*** | -1.347*** | -1.347*** |
| | [-10.70] | [-10.70] | [-10.70] | [-10.86] | [-10.86] | [-10.87] |
| Memorial day | -2.512*** | -2.512*** | -2.512*** | -2.517*** | -2.517*** | -2.517*** |
| | [-17.64] | [-17.64] | [-17.64] | [-17.65] | [-17.66] | [-17.65] |
| N | 185671 | 185671 | 185671 | 185671 | 185671 | 185671 |
| R-sq | 0.716 | 0.716 | 0.716 | 0.717 | 0.717 | 0.717 |
| adj. R-sq | 0.716 | 0.716 | 0.715 | 0.716 | 0.716 | 0.716 |
| AIC | 1023156.9 | 1023158.0 | 1023168.6 | 1022790.1 | 1022795.1 | 1022801.2 |
| BIC | 1023643.2 | 1023624.1 | 1023614.4 | 1023276.4 | 1023261.2 | 1023246.9 |

t statistics in brackets * p<0.10, ** p<0.05, *** p<0.01

Note: Holidays of the second half of the year are out of sample and their dummy variables removed

**Table S9. Station Fixed effect models using data from January to June, 2019 to 2020 with different post period specifications**

| | P1 & P2 | | | P2 & P3 | | |
|---|---|---|---|---|---|---|
| | NO2 | NO2 | NO2 | NO2 | NO2 | NO2 |
| P1 | -1.454*** | -1.479*** | -1.795*** | | | |
| | [-3.84] | [-3.89] | [-4.75] | | | |
| P2 | -3.6*** | -3.617*** | -3.716*** | -5.065*** | -5.087*** | -5.148*** |
| | [-11.20] | [-11.39] | [-12.02] | [-13.08] | [-13.29] | [-14.46] |
| P3 | | | | -3.512*** | -3.534*** | -3.442*** |
| | | | | [-7.45] | [-7.56] | [-8.37] |
| P1*RepG | .5878*** | .4649*** | | | | |
| | [3.34] | [2.74] | | | | |
| P2*RepG | .2147 | .1447 | | .1742 | .0872 | |
| | [1.06] | [0.82] | | [0.70] | [0.40] | |
| P3*RepG | | | | -.0433 | -.144 | |
| | | | | [-0.17] | [-0.62] | |
| P1*TrumpC | -.8301* | | | | | |
| | [-1.90] | | | | | |
| P2*TrumpC | -.467 | | | -.5802 | | |
| | [-0.99] | | | [-1.02] | | |
| P3*TrumpC | | | | -.6718 | | |
| | | | | [-1.17] | | |
| P1*TrumpS | 4.419*** | 3.701*** | 4.847*** | | | |
| | [4.45] | [4.41] | [6.27] | | | |
| P2*TrumpS | 6.419*** | 6.024*** | 6.381*** | 7.27*** | 6.78*** | 6.999*** |
| | [8.23] | [8.40] | [10.24] | [7.77] | [7.86] | [10.17] |
| P3*TrumpS | | | | 4.478*** | 3.907*** | 3.566*** |
| | | | | [4.13] | [3.72] | [4.62] |
| Precipitation in mm | -.0102 | -.0101 | -.0102 | -.0105 | -.0107 | -.0107 |
| | [-1.32] | [-1.32] | [-1.32] | [-1.36] | [-1.39] | [-1.39] |
| Precipitation in mm ^2 | 4.1e-04*** | 4.1e-04*** | 4.1e-04*** | 4.1e-04*** | 4.2e-04*** | 4.2e-04*** |
| | [3.24] | [3.24] | [3.24] | [3.30] | [3.31] | [3.31] |
| Dew point in Celsius | -.1732*** | -.1732*** | -.173*** | -.1709*** | -.1704*** | -.1701*** |
| | [-11.14] | [-11.14] | [-11.13] | [-11.11] | [-11.18] | [-11.11] |
| Dew point in Celsius ^2 | -.0018*** | -.0018*** | -.0018*** | -.0018*** | -.0018*** | -.0018*** |
| | [-4.23] | [-4.28] | [-4.27] | [-4.21] | [-4.21] | [-4.20] |
| Pressure in Pa | .0034 | .0033 | .0032 | .0045 | .0045 | .0046 |
| | [0.58] | [0.56] | [0.54] | [0.78] | [0.78] | [0.79] |
| Pressure in Pa ^2 | -1.6e-08 | -1.5e-08 | -1.5e-08 | -2.1e-08 | -2.1e-08 | -2.1e-08 |
| | [-0.54] | [-0.52] | [-0.50] | [-0.74] | [-0.74] | [-0.75] |
| Wind intensity in m/s | -2.881*** | -2.882*** | -2.882*** | -2.859*** | -2.862*** | -2.862*** |
| | [-24.05] | [-24.02] | [-24.02] | [-23.92] | [-23.88] | [-23.89] |
| Wind intensity in m/s ^2 | .1815*** | .1816*** | .1816*** | .1794*** | .1797*** | .1798*** |
| | [15.28] | [15.27] | [15.28] | [15.13] | [15.12] | [15.12] |
| Temperature in Celsius | .1645*** | .1646*** | .1644*** | .1646*** | .1635*** | .1632*** |
| | [9.54] | [9.56] | [9.54] | [9.60] | [9.61] | [9.55] |
| Saturday | -1.416*** | -1.416*** | -1.416*** | -1.412*** | -1.412*** | -1.412*** |
| | [-18.56] | [-18.56] | [-18.56] | [-18.48] | [-18.48] | [-18.48] |
| Sunday | -2.191*** | -2.191*** | -2.191*** | -2.201*** | -2.201*** | -2.201*** |
| | [-22.29] | [-22.29] | [-22.29] | [-22.39] | [-22.39] | [-22.39] |
| New Year's Day | -3.609*** | -3.609*** | -3.609*** | -3.599*** | -3.599*** | -3.599*** |
| | [-19.24] | [-19.24] | [-19.24] | [-19.22] | [-19.23] | [-19.22] |
| Martin Luther King Jr. Day | -1.413*** | -1.413*** | -1.413*** | -1.409*** | -1.411*** | -1.411*** |
| | [-8.66] | [-8.66] | [-8.66] | [-8.68] | [-8.69] | [-8.69] |
| Presidents day | -1.366*** | -1.365*** | -1.365*** | -1.399*** | -1.399*** | -1.4*** |
| | [-10.82] | [-10.83] | [-10.83] | [-10.99] | [-10.98] | [-10.98] |
| Memorial day | -2.347*** | -2.347*** | -2.347*** | -2.353*** | -2.353*** | -2.352*** |
| | [-17.74] | [-17.74] | [-17.74] | [-17.77] | [-17.77] | [-17.76] |
| N | 124418 | 124418 | 124418 | 124418 | 124418 | 124418 |
| R-sq | 0.714 | 0.713 | 0.713 | 0.714 | 0.714 | 0.714 |
| adj. R-sq | 0.713 | 0.713 | 0.713 | 0.713 | 0.713 | 0.713 |
| AIC | 676489.8 | 676497.7 | 676510.0 | 676127.8 | 676146.6 | 676149.7 |
| BIC | 676898.5 | 676887.0 | 676879.8 | 676536.5 | 676535.8 | 676519.5 |

t statistics in brackets * p<0.10, ** p<0.05, *** p<0.01

Note: Holidays of the second half of the year are out of sample and their dummy variables removed

STATA CODE:

```
***********************************************************************
cd "..working directory"



**********************************************************************
********** Merging data sets         ****************************************
**********************************************************************

use polutant_total_est.dta,clear
replace latit=round( latitude, 0.001)
replace longit=round(longitude,0.001)
merge m:1 statecode countycode sitenum using stationsUSA.dta
drop if _merge!=3
drop _merge
merge m:m statecode countycode id date using meteoUSA.dta
drop if _merge!=3
drop _merge
merge m:m statecode using elections_county.dta
drop _merge
merge m:m statecode date using epidemy_state.dta

rename _merge merge_3
replace merge_3=3 if date<21937
replace newc_per100k=0 if date<21937
replace newd_per100k =0  if date<21937
replace totc_per100k =0  if date<21937
replace totd_per100k=0  if date<21937
drop if merge_3!=3    /*** drops District of Columbia and post 06/30/2020 observations
***/
drop merge_3
drop if dewt==0 & pres==0 /* drop Alaska and Hawaii  */




**********************************************************************
********** Moving averages *********************************************
**********************************************************************

tsset id date
tssmooth ma no_ma =NO2, window(364 1 0)
```

```
tssmooth ma no_mw =NO2, window(6 1 0)
tssmooth ma no_mm =NO2, window(27 1 0)



************************************************************************
********** New variables ***********************************************
************************************************************************

***************  Date variables ******************************

gen year=year(date)
gen month=month(date)
gen sunday=  dow(date)==0
gen saturday=dow(date)==6
gen quarter= quarter(date)
gen week=    week(date)


**** meteo squared variables   ******************************
gen pres2=pres^2
gen dewt2=dewt^2
gen wpow2=wpow^2
gen tpre2=tpre^2


************* Federal holidays variables *********************

gen new_year=0
replace new_year= 1 if (month(date) == 1 & day(date) == 1)

gen mlk=0
replace mlk = 1 if (month(date) ==1 & day(date) == 15  & year==2018)
replace mlk = 1 if (month(date) ==1 & day(date) == 21  & year==2019)
replace mlk = 1 if (month(date) ==1 & day(date) == 20  & year==2020)

gen president=0
replace president = 1 if (month(date) ==2 & day(date) == 19 & year==2018)
replace president = 1 if (month(date) ==2 & day(date) == 18 & year==2019)
replace president = 1 if (month(date) ==2 & day(date) == 17 & year==2020)

gen memorial=0
replace memorial = 1 if (month(date) ==5 & day(date) == 28  & year==2018)
replace memorial = 1 if (month(date) ==5 & day(date) == 27  & year==2019)
replace memorial = 1 if (month(date) ==5 & day(date) == 25  & year==2020)
```

```
gen fourth_july=0
replace fourth_july = 1 if (month(date) ==7 & day(date) == 4)

gen labor=0
replace labor = 1 if (month(date) ==9 & day(date) == 3 & year==2018)
replace labor = 1 if (month(date) ==9 & day(date) == 2 & year==2019)

gen colombus=0
replace colombus = 1 if (month(date) ==10 & day(date) == 8  & year==2018)
replace colombus = 1 if (month(date) ==10 & day(date) == 14 & year==2019)


gen veteran=0
replace veteran = 1 if (month(date) ==11 & day(date) == 12)
replace veteran = 1 if (month(date) ==11 & day(date) == 11 & year==2019)

gen thanks_giving=0
replace thanks_giving = 1 if (month(date) ==11 & day(date) == 22 & year==2018)
replace thanks_giving = 1 if (month(date) ==11 & day(date) == 28 & year==2019)

gen da_thanks_giving=0
replace da_thanks_giving = 1 if (month(date) ==11 & day(date) == 23 & year==2018)
replace da_thanks_giving = 1 if (month(date) ==11 & day(date) == 29 & year==2019)

gen christmas=0
replace christmas = 1 if (month(date) == 12 & day(date) == 25)


*****************   region variables  ******************
gen south= latit<=35.15
gen north= latit>=40.25
gen center_ns= south==0 & north==0
gen lat2=2
replace lat2=1 if south==1
replace lat2=3 if north==1

gen east= longit<=-104.75
gen west= longit>=-84.25
gen center_ew= (east==0 & west==0)
gen longi2=2
replace longi2=1 if east==1
replace longi2=3 if west==1

gen region=(3*longi2+lat2)-3
```

```
*************** post subperiod variables
********************************************

gen post1=    date>=21971 & date<21987          /* 26th Feb, First comunity case*/
gen post2=    date>=21987 & date<22019          /* National emergency declaration */
gen post3=    date>=22019          /* trasfer to the States 16 de abril */


**************** did variables
****************************************************

gen did1=post1*republic
gen did2=post2*republic
gen did3=post3*republic

gen did1c_t16=post1*perc_c_trump_16
gen did2c_t16=post2*perc_c_trump_16
gen did3c_t16=post3*perc_c_trump_16

gen did1s_t16=post1*perc_trump_16
gen did2s_t16=post2*perc_trump_16
gen did3s_t16=post3*perc_trump_16

save science_adv.dta, replace



************************************************************************
********** Statition fixed effects models *****************************
************************************************************************
use science_adv.dta, clear

global t1 (year>=2018)
global t2 (year>=2019)
global t3 (year>=2018 & month<7)
global t4 (year>=2019 & month<7)



global m i.reg#quarter tpre tpre2 dewt dewt2 pres pres2 wpow wpow2 temp
global h new_year mlk president memorial fourth_july labor colombus veteran
thanks_giving da_thanks_giving christmas

global t13  did1-did3
global tc13 did1c_t16-did3c_t16
global ts13 did1s_t16-did3s_t16
```

```
global t12  did1-did2
global tc12 did1c_t16-did2c_t16
global ts12 did1s_t16-did2s_t16


global t23  did2-did3
global tc23 did2c_t16-did3c_t16
global ts23 did2s_t16-did3s_t16



foreach t in t1 t2 t3 t4  {
areg NO2 year#month republic post1-post3  $t13 $tc13 $ts13 $m saturday sunday $h if
$`t', vce(cluster id) ab(id)
estimates store FE_NO2_GCS_post_1_3_`t'
areg NO2 year#month republic post1-post3  $t13      $ts13 $m saturday sunday $h if $`t',
vce(cluster id) ab(id)
estimates store FE_NO2_GS_post_1_3_`t'
areg NO2 year#month republic post1-post3  $t13 $tc13      $m saturday sunday $h if $`t',
vce(cluster id) ab(id)
estimates store FE_NO2_GC_post_1_3_`t'
areg NO2 year#month republic post1-post3      $tc13 $ts13 $m saturday sunday $h if $`t',
vce(cluster id) ab(id)
estimates store FE_NO2_CS_post_1_3_`t'
areg NO2 year#month republic post1-post3  $t13           $m saturday sunday $h if $`t',
vce(cluster id) ab(id)
estimates store FE_NO2_G_post_1_3_`t'
areg NO2 year#month republic post1-post3      $tc13       $m saturday sunday $h if $`t',
vce(cluster id) ab(id)
estimates store FE_NO2_C_post_1_3_`t'
areg NO2 year#month republic post1-post3           $ts13 $m saturday sunday $h if $`t',
vce(cluster id) ab(id)
estimates store FE_NO2_S_post_1_3_`t'


areg NO2 year#month republic post1-post2  $t12 $tc12 $ts12 $m saturday sunday $h if
$`t', vce(cluster id) ab(id)
estimates store FE_NO2_GCS_post_1_2_`t'
areg NO2 year#month republic post1-post2  $t12       $ts12 $m saturday sunday $h if $`t',
vce(cluster id) ab(id)
estimates store FE_NO2_GS_post_1_2_`t'
areg NO2 year#month republic post1-post2  $t12 $tc12       $m saturday sunday $h if $`t',
vce(cluster id) ab(id)
estimates store FE_NO2_GC_post_1_2_`t'
```

```
areg NO2 year#month republic post1-post2       $tc12 $ts12 $m saturday sunday $h if $`t', vce(cluster id) ab(id)
estimates store FE_NO2_CS_post_1_2_`t'
areg NO2 year#month republic post1-post2  $t12            $m saturday sunday $h if $`t', vce(cluster id) ab(id)
estimates store FE_NO2_G_post_1_2_`t'
areg NO2 year#month republic post1-post2      $tc12        $m saturday sunday $h if $`t', vce(cluster id) ab(id)
estimates store FE_NO2_C_post_1_2_`t'
areg NO2 year#month republic post1-post2            $ts12  $m saturday sunday $h if $`t', vce(cluster id) ab(id)
estimates store FE_NO2_S_post_1_2_`t'


areg NO2 year#month republic post2-post3  $t23 $tc23 $ts23 $m saturday sunday $h if $`t', vce(cluster id) ab(id)
estimates store FE_NO2_GCS_post_2_3_`t'
areg NO2 year#month republic post2-post3  $t23       $ts23 $m saturday sunday $h if $`t', vce(cluster id) ab(id)
estimates store FE_NO2_GS_post_2_3_`t'
areg NO2 year#month republic post2-post3  $t23 $tc23       $m saturday sunday $h if $`t', vce(cluster id) ab(id)
estimates store FE_NO2_GC_post_2_3_`t'
areg NO2 year#month republic post2-post3       $tc23 $ts23 $m saturday sunday $h if $`t', vce(cluster id) ab(id)
estimates store FE_NO2_CS_post_2_3_`t'
areg NO2 year#month republic post2-post3  $t23            $m saturday sunday $h if $`t', vce(cluster id) ab(id)
estimates store FE_NO2_G_post_2_3_`t'
areg NO2 year#month republic post2-post3      $tc23        $m saturday sunday $h if $`t', vce(cluster id) ab(id)
estimates store FE_NO2_C_post_2_3_`t'
areg NO2 year#month republic post2-post3            $ts23  $m saturday sunday $h if $`t', vce(cluster id) ab(id)
estimates store FE_NO2_S_post_2_3_`t'

}




****************************************************************************
********** TABLE RESULTS  CSV        *************
****************************************************************************
```

```
esttab * using science_advno22.csv, replace scsv bracket b(%6.4g) t(2) keep(republic
post* did* saturday sunday $h tpre tpre2 dewt dewt2 pres pres2 wpow wpow2 temp) r2
ar2 aic(%10.1f)  bic(%10.1f)   star(* 0.10 ** 0.05 *** 0.01)
estimates clear



*************************************************************************
********** Fixed effects per station and subperiods models ************
*************************************************************************


estimates clear
forvalues i = 1(1)5 {
reg NO2 year#month i.id i.post1#i.id i.post2#i.id i.post3#i.id saturday sunday $m $h if
$t1, vce(cluster id)
estimates store FE_NO2_postid_t1
reg NO2 year#month i.id i.post1#i.id i.post2#i.id i.post3#i.id saturday sunday $m $h if
$t2, vce(cluster id)
estimates store FE_NO2_postid_t2
reg NO2 year#month i.id i.post1#i.id i.post2#i.id i.post3#i.id saturday sunday $m $h if
$t3, vce(cluster id)
estimates store FE_NO2_postid_t3
reg NO2 year#month i.id i.post1#i.id i.post2#i.id i.post3#i.id saturday sunday $m $h if
$t4, vce(cluster id)
estimates store FE_NO2_postid_t4

}

**************************************************************************
********** TABLE STATION FIXED EFFECTS PER SUBPERIOD
*************
**************************************************************************
esttab * using fe_postid_table, replace scsv bracket keep(1.post1#* 1.post2#* 1.post3#*)
nostar wide nopa



********************************************************************
********* FIGURES       *******************************************
********************************************************************

*********** FIGURE 2  ****************************************
use science_adv.dta, clear
```

```
collapse (mean) NO2 *_m* year month saturday sunday $h reg quarter tpre tpre2 dewt
dewt2 pres pres2 wpow wpow2 temp did* post*, by(date)


twoway (line no_mm date , lcolor(red)) if date>21200, xline(21971, lcolor(gs10)
lpattern(dash))  ytitle(Average NO2 emissions) xtitle(Date) scheme(s2color) legend(
size(vsmall)) text(12 21970 "First community", place(e) just(left) size(vsmall)) text(11.75
21970 "spread case", place(e) just(left) size(vsmall))
graph save Fig2, replace
graph export "Fig2.pdf", replace as(pdf)



*********** FIGURES 1 &  3 ****************************************
use science_adv.dta, clear
collapse (mean) NO2 *_m* year month saturday sunday $h reg quarter tpre tpre2 dewt
dewt2 pres pres2 wpow wpow2 temp did* post*, by(republic date)
sort date republic

gen difNO2= NO2 - NO2[_n-1] if date==date[_n-1] & republic==1

tsset republic date
tssmooth ma difno_w =difNO2, window(55 1 0)

twoway (line difno_w date if date>=21239,  lcolor(red)) , ytitle(Average differences in
NO2 emissions) scheme(s2color) xline(22033 21302 21667, lw(12) lcolor(gs14)) yline(0,
lcolor(gs10)) xline(21987, lcolor(blue))
graph save Fig3, replace
graph export "Fig3.pdf", replace as(pdf)


twoway (line no_mw date if republic==1, lcolor(red))  (line no_mw date if republic==0,
lcolor(blue)) if year==2020, xline(21971 21987 22019, lcolor(gs10) lpattern(dash))
ytitle(Average NO2 emissions) xtitle(Date) scheme(s2color) legend( size(vsmall) lab(1
"Republican") lab(2 "Democratic")) text(14 21958 "First community", place(e) just(left)
size(vsmall)) text(13.75  21960.75 "spread case", place(e) just(left) size(vsmall))
text(13.25 21972 "National emergency", place(e) just(left) size(vsmall)) text(13  21978
"declaration", place(e) just(left) size(vsmall))  text(14  22009 "Trump transfers total",
place(e) just(left) size(vsmall)) text(13.75 22011.25 "authority to states", place(e)
just(left) size(vsmall))
graph save Fig1, replace
graph export "Fig1.pdf", replace as(pdf)
```
